# A phase 1 randomized, placebo-controlled trial of a combination typhoid and non-typhoidal *Salmonella* polysaccharide conjugate vaccine

**DOI:** 10.1101/2025.09.15.25335795

**Authors:** Wilbur H. Chen, Robin S. Barnes, Michael J. Sikorski, Reva Datar, Roohali Sukhavasi, Yuanyuan Liang, Rekha R. Rapaka, Marcela F. Pasetti, Marcelo B. Sztein, Rezwanul Wahid, Sharon M. Tennant, Raphael Simon, Scott M. Baliban, James E. Galen, Andrew Lees, Biana Bernshtein, Galit Alter, Raches Ella, Krishna Mohan, M. Gangadhara Naidu, D. Yogeswar Rao, Krishna M. Ella, Myron M. Levine

**Affiliations:** Center for Vaccine Development and Global Health, University of Maryland School of Medicine, Baltimore, MD, U.S.A; Department of Epidemiology and Public Health, University of Maryland School of Medicine, Baltimore, MD, U.S.A; Fina Biosolutions, LLC, Rockville, MD, U.S.A; Ragon Institute, Harvard Medical School, Cambridge, MA, U.S.A; Bharat Biotech International Ltd, Hyderabad, Telangana, India

## Abstract

In sub-Saharan Africa, non-typhoidal *Salmonella* (NTS) and *Salmonella* Typhi are leading causes of invasive disease among young children. Trivalent *Salmonella* Conjugate Vaccine (TSCV) consists of Vi capsule polysaccharide conjugated to tetanus toxoid and core-plus-O-polysaccharides from the two most prevalent invasive non-typhoidal serovars (Typhimurium, Enteritidis) conjugated to serovar- homologous flagellin subunits. We conducted a first-in-human, randomized, placebo-controlled, stepwise-dose-escalation phase 1 trial (NCT03981952) evaluating TSCV safety and immunogenicity; 22 healthy adults aged 18-45 years were randomly allocated 6.25 µg TSCV (n=8), 12.5 µg TSCV (n=10), or placebo (n=4). TSCV was safe and well-tolerated, with the most common solicited symptom being short- lived injection site pain. For each of the 3 polysaccharides, serum IgG and IgA ELISA antibody responses, as demonstrated by four-fold or greater increases over baseline, were observed among all vaccinees but among no placebo recipients. Binding and functional antibodies, gut-homing antibody secreting cells, and polysaccharide-specific memory B cells responses were also elicited.

ClinicalTrials.gov Registry NCT03981952.

## Introduction

In industrialized countries, typhoid fever, caused by *Salmonella enterica* serovar Typhi, is rare (except among travelers) and non-typhoidal *Salmonella* (NTS) infections, which are common, typically manifest as self-limited enterocolitis. Whereas invasive clinical syndromes caused by NTS serovars are rare in industrialized countries, in young children of sub-Saharan Africa (SSA), febrile bacteremia, septicemia, and meningitis caused by invasive NTS (iNTS) are common.^1–5^ In 2017 it was estimated that globally ∼535,000 cases of iNTS disease occurred in persons not infected with HIV, of which ∼421,600 transpired in SSA, accompanied by ∼66,520 deaths.^6^ This estimated morbidity and mortality burden in SSA mainly befell children <5 years of age.

Among the many possible NTS serovars, *Salmonella enterica* serovar Typhimurium and other Group O:4 (B) serovars and *Salmonella enterica* serovar Enteritidis and other Group O:9 (D) serovars are collectively responsible for almost 90% of the young child cases of iNTS.^5^ Also, circa 80% of all iNTS cases are in children <5 years of age with infants in the first semester of life having a markedly lower burden than infants 6-11 months of age. Thus, the magnitude and age-specific burden of iNTS disease in SSA closely resembles that of invasive *Haemophilus influenzae* type b (Hib) disease before the introduction of Hib conjugate vaccine (as a component of pentavalent vaccine) in African countries.^7^ Collectively, these data suggest that if a safe and efficacious iNTS vaccine can be developed that is readily deliverable to infants, and if high coverage can be achieved through the Essential Programme on Immunization (EPI), the burden of severe and fatal iNTS infections in African infants and toddlers can be markedly reduced, as occurred with invasive Hib disease.^7^

Typhoid fever is also a public health burden in many regions of SSA, albeit more so among older pre-school and school age children. Typhoid was estimated to have caused 653,200 cases and 8,751 deaths in Western sub-Saharan Africa in 2017.^8^ *S*. Paratyphi A disease is rare in SSA. It is not uncommon in SSA for iNTS or typhoid to predominate in different regions of the same country. Since 2018, Vi conjugate typhoid vaccine has been introduced into multiple countries in SSA, some of which also have known burdens of iNTS disease.^9^

It is thus advantageous to consider a Trivalent *Salmonella* Conjugate Vaccine (TSCV) that would confer protection against both typhoid and iNTS disease. Accordingly, the Center for Vaccine Development and Global Health (CVD) of the University of Maryland School of Medicine and Bharat Biotech International Limited (BBIL) have jointly designed and constructed a TSCV in which the core and O-polysaccharide (COPS) of *S*. Typhimurium and of *S*. Enteritidis are individually covalently linked to the respective Phase 1 flagellin subunits of the homologous serovar^10,11^ and are formulated in combination with *S*. Typhi Vi conjugate Typbar TCV™,^12^ the first Vi conjugate to be prequalified by the World Health Organization^13^ and approved for support from GAVI – the Vaccine Alliance.^14^

One notable innovation of the iNTS components of the CVD/BBIL TSCV is use of a carrier protein derived from the homologous serovar such that the carrier serves as a protective antigen in addition to the COPS antigen and provides T-helper epitopes from the target pathogens.^10,11,15,16^ Another innovation is that the iNTS COPS and FliC antigens derive from engineered “reagent strains” that utilize optimized OPS-chain length for enhanced immunogenicity and hyper-express FliC subunits consequent to the mutation in *clpXP*.^17^ Moreover, consequent to the deletion of *fliD* the hyper-expressed FliC monomers are exported directly into the fermentation medium, thereby facilitating antigen purification.^17^ Preclinical studies in mice and rabbits demonstrated robust immunogenicity to the respective COPS and flagellin antigens after immunization with the iNTS conjugates. Mice that were actively immunized with the COPS:FliC conjugates were also significantly protected against homologous serovar challenge with iNTS invasive clinical isolates representing the dominant circulating multi-locus sequence types in SSA.

Based on the supportive preclinical data, and the demonstration of TSCV’s safety and immunogenicity in a Rabbit Toxicology Study under Good Laboratory Practices,^12^ with approval of the clinical protocol by the UMB Institutional Review Board, TSCV was cleared to enter a Phase 1 randomized, double-blinded, placebo-controlled, stepwise dose-escalation clinical trial under regulatory oversight of the U.S. Food and Drug Administration. The primary objectives of the Phase 1 study was to establish safety and immunogenicity, based on the elicitation of antigen-specific serum IgG antibody.

## Methods

### Vaccine and placebo

The Trivalent *Salmonella* Conjugate Vaccine (TSCV) was manufactured by Bharat Biotech International Limited (BBIL, Hyderabad, India) and includes three conjugate components. The *S*. Enteritidis conjugate component was generated by 1-cyano-4-dimethylaminopyridinium tetrafluoroborate (CDAP) chemistry to link *S*. Enteritidis core O-polysaccharide (COPS) to the homologous serovar FliC phase 1 flagellin protein subunit (FliC) that had been derivatized with adipic acid dihydrazide (ADH) using carbodiimide. The *S*. Typhimurium conjugate component was generated by modification of the polysaccharide terminal 2-keto-3-deoxyoctonate (KDO) carbonyl with an aminooxy thiol linker. The aminooxy forms an oxime bond with the KDO ketone that was further reduced with sodium cyanoborohydride. *S*. Typhimurium FliC protein was derivatized with the amine reactive reagent succinimidyl 4-maleimidylbutyrate (GMBS) to introduce a maleimide moiety that was then linked to the reactive sulfhydryl of the derivatized COPS molecule via formation of a thioether bond. The Typhoid Vi conjugate component was essentially the BBIL Typbar-TCV™ vaccine which is WHO pre-qualified and licensed in India. Typbar-TCV™ is generated by carbodiimide mediated modification of *S*. Typhi Vi capsular polysaccharide (CPS) with ADH that introduces reactive hydrazide groups that are then used to link to tetanus toxoid (TT) via a second carbodiimide step.

The TSCV was formulated in multi-dose vials as a 1:1:1 mixture of the three conjugate components in sterile buffered saline (PBS) with 0.02% Tween-80, pH 7.2, plus 10 mg/mL of 2- Phenoxyethanol. Each vial held material for which a 0.5 mL volume contained 25 μg of each of the three respective polysaccharides by weight. The placebo was formulated by BBIL in similar multi-dose vials containing only the buffer vehicle in which the TSCV was suspended, plus 10 mg/mL of 2- Phenoxyethanol. A 6.25 μg (of each polysaccharide by weight) dose of vaccine was prepared by mixing a 0.25 mL volume from the TSCV vial with 0.75 mL of placebo and administering a 0.5 mL volume of this dilution. A 12.5 μg dose of vaccine was prepared by mixing 0.5 mL from the TSCV vial with 0.5 mL of placebo and administering a 0.5 mL volume of this dilution.

### Study Design and participants

This first-in-human, randomized, double-blinded, placebo-controlled, phase 1 trial enrolled a total of 22 adult male and non-pregnant female participants, aged 18-45 years. Participants were excluded if they had a history of a clinically significant comorbid disease or prior history of typhoid infection or vaccination within the prior 5 years. The detailed inclusion and exclusion criteria are described in the Study Protocol. Prior to randomization and receipt of blinded study vaccine, eligibility criteria were confirmed. Following a single 0.5 mL intramuscular dose of blinded study vaccine, participants were monitored for 20 minutes. The trial was conducted at the Center for Vaccine Development and Global Health, Baltimore, Maryland, U.S.A. The protocol was approved by the University of Maryland Baltimore Institutional Review Board and all participants provided written informed consent. ClinicalTrials.gov identifier NCT03981952.

An initial cohort (Cohort A) of 10 participants were enrolled 09-11 December 2019 and consisted of 8 participants randomized to receive 6.25 μg vaccine and 2 participants to placebo. After an independent safety monitoring committee reviewed the available safety data through at least 7-days post-vaccination, a following cohort (Cohort B) of 12 participants were enrolled 18-20 February 2020 and consisted of 10 participants randomized to receive 12.5 μg vaccine and 2 participants to placebo.

Meanwhile, the emergence of the SARS-CoV-2 virus caused a U. S. Public Health Emergency declaration (since 27 January 2020) and local site research restrictions which began 19 March 2020. A planned evaluation of a 25 μg dose of vaccine were unable to be conducted due to the ongoing pandemic restrictions and subsequent expiration of the study vaccine. Local site pandemic closures prevented in-person follow up visits at 6-months for all participants of Cohort A and 2-months and 6-months for all participants of Cohort B. There were also 2 participants in Cohort B who did not complete the 28-day post-vaccination visit and thus are excluded from the immunogenicity endpoint analysis. However, a supplemental in-person visit was completed at approximately 450-510 days post-vaccination, for the evaluation of longer-term immunogenicity, by 16 of the 20 participants included in the immunogenicity endpoint analysis.

### Outcomes

The primary objective of this study was to evaluate the safety and tolerability of escalating doses of TSCV. A co-primary immunogenicity objective was assessed through the assessment of seroconversions in vaccine antigen-specific serum IgG ELISA titers, at 28-days and 57-days post-vaccination. A secondary safety objective consisted of the measurement of clinical safety laboratory parameters at 7 days post-vaccination. The secondary immunogenicity objective was the measurement of vaccine antigen-specific serum IgG ELISA responses at other timepoints. Exploratory immunogenicity objectives included the evaluation of vaccine antigen-specific antibody secreting cells (ASCs) and their homing markers, antibodies in lymphocyte supernatants (ALS), memory B and T cells, and antibody profiling.

### Safety Evaluation

Following receipt of blinded study product, participants were provided diaries to record local or systemic solicited adverse events (AEs) over 7 days. Solicited local symptoms included injection site pain, erythema, induration, and ecchymosis. Solicited systemic symptoms included fever, chills, fatigue, malaise, myalgia, arthralgia, nausea, and headache. Unsolicited AEs were collected over 28 days and serious adverse events (SAEs) over 6 months. Because of the pandemic, the protocol was modified, and SAEs were collected over 450-510 days (∼510 days for Cohort A and ∼450 days for Cohort B), for some individuals. Clinical laboratories measured at baseline and 7-days post-vaccination included total white blood cells count, absolute neutrophil count, hemoglobin, platelet count, creatinine, alanine aminotransferase, aspartate aminotransferase, and total bilirubin. The C-Reactive Protein (CRP) level was measured at baseline, 1-day and 7-days post-vaccination.

### ELISA

The serum IgG and IgA levels against the respective antigens were measured, as previously described.^1–3^ Briefly, 96-well plates were coated with the respective COPS, Vi, or FliC antigen (5 µg/mL). After overnight incubation, plates were washed and sera were tested in serial dilutions. Specific antibodies were detected using peroxidase-labeled anti-human IgG and 3,3,5,5-Tetramethylbenzidine (TMB) substrate. Test and control sera were run in duplicate. Titers were calculated by interpolation of absorbance values of test samples into the linear regression curve of a calibrated control (reference serum). The endpoint titers are reported as ELISA units (EU), which represented the inverse of the serum dilution that produced an absorbance value of 0.2 above the blank. Seroconversion was defined as a 4- fold increase in the antibody titer after immunization.

### ASC assays

A standard B cell ELISPOT assay, as previously described,^4–7^ was used to measure ASC in freshly purified PBMC collected from volunteers participating in Cohort B (12.5 µg dose group, Vaccinees; n=10, Placebo; n=2). Briefly, duplicate wells of MAHA Plates (Millipore, MAHAS4510) were coated with 1. Vi-biotin (5 µg/mL); 2. Lipopolysaccharide (LPS) from *S*. Enteritidis (Sigma#L6011) and *S*. Typhimurium (Sigma#L6511); 3. Flagellin (FliC, 5 µg/mL) prepared in-house from *S*. Enteritidis (SE) and *S*. Typhimurium (STm); and 4. Tetanus Toxoid (TT, Connaught Laboratories, Toronto, Canada). Freshly purified PBMC (200,000 cells/well for each antigen) were added to each well and incubated overnight (O/N). The next day, spots were developed and visualized using HRP-conjugated IgG and IgA secondary antibodies and 3-Amino-9-ethylcarbazole (AEC) substrate. Plates were read and QC’d using an automated reader (Cellular Technology Limited, CTL, Cleveland, Ohio) and CTL software. When there were too many spots to count/well, counts were capped at 175 spots/well. Data were calculated as spot-forming cells (SFC)/10^6^ PBMC based on the number of cells seeded/well (200,000 cells).

### B cell sorting

We utilized an optimized flow cytometry-based sorting protocol to simultaneously sort 5 subpopulations of B cells based on their surface expression of different memory and homing markers as previously described.^8^ Using this protocol, we sorted B cells (live CD3-CD19+) subpopulations based on their expression of the memory marker CD27 into naïve B cells (CD27-) and memory (CD27+) B cells.

Memory B cells were sorted into 4 different subsets based on their expression of integrin α4β7 and CD62L homing molecules, e.g., CD3-CD19+CD27+integrin α4β7-CD62L+ (62L Single Positives; 62L SP), CD3-CD19+CD27+integrin α4β7-CD62L- (Double Negatives; DN), CD3-CD19+CD27+integrin α4β7+CD62L+ (Double Positives; DP) and CD3-CD19+CD27+integrin α4β7+CD62L- (α4 SP) subsets as shown in **Supplementary Figure 7**. These highly purified subsets of B cells were evaluated for their ability to secrete specific antibodies in an antigen-specific ELISPOT assay as described above for ASC. Each subpopulation was distributed into each of duplicate wells per antigen evaluated with a minimum 1,000 cells seeded per well. The average SFC measured in duplicate antigen coated wells were converted to SFC/10^6^ cells based on the number of seeded cells/well for each subset.

Flow cytometry measurements and sorting were performed with MoFlow Astrios cell sorter (Beckman- Counter) at the Flow Cytometry and Mass Cytometry Core Facility of the University of Maryland School of Medicine Center for Innovative Biomedical Resources (CIBR), Baltimore, Maryland. Samples were analyzed using a FlowJo software package (Tree Star, USA).

### B memory assays

Antigen-specific B memory cells were performed as we previously reported.^6,9–11^ Briefly, thawed PBMC were stimulated in vitro with a B cell-expansion media consisting of 5 μM 2β-ME (Biorad), 1:100,000 pokeweed mitogen (kindly provided by Dr. S. Crotty), 6 μg/mL CpG-2006 (Qiagen/Operon, Huntsville, AL), and 1:10,000 *Staphylococcus aureus* Cowan (SAC, Sigma–Aldrich, St. Louis, MO) for 5 days. Antigen-specific B ELISPOT assays were performed as described for ASC assays with these expanded cells. In addition to the antigens mentioned for ASC assays above, cells were also seeded for capturing non-specific (total) IgG and IgA secreting BM cells. Specific spot forming cells (SFC) were converted to SFC/10^6^ of expanded cells, according to the number of cells seeded/well as described above. The IgG and IgA BM data are presented as the percentages of the corresponding non-specific or total IgG or IgA (i.e., % of Total) BM.

### Luminex-based antibody profiling

Antigen-specific antibody subclass and isotype binding levels were assessed using a 384-well based customized multiplexed Luminex assay, as previously described.^12,13^ *S.* Typhi Vi, *Citrobacter freundii* Vi, *S*. Enteritidis COPS, *S*. Typhimurium COPS, *S*. Enteritidis FliC, *S*.Typhimurium FliC, and *S*. Choleraesuis LPS were used to profile the antigen-specific humoral immune response. Tetanus toxin and Ebola (CEFTA, Mabtech Inc.) were used as control antigens. Protein antigens were coupled to magnetic Luminex beads (Luminex Corp) by carbodiimide-NHS ester-coupling (Thermo Fisher). COPS and LPS were coupled to magnetic Luminex beads (Luminex Corp) by carbodiimide-NHS ester-coupling (Thermo Fisher). Antigens were modified by 4-(4,6-dimethoxy[1,3,5]triazin-2-yl)-4-methyl-morpholinium and conjugated to Luminex Magplex carboxylated beads. Antigen-coupled microspheres were washed and incubated with serum samples at an appropriate sample dilution (1:200) for 2 hours at 37°C in 384-well plates (Greiner Bio-One). Unbound antibodies were washed away, and antigen-bound antibodies were detected by using a PE-coupled detection antibody for each subclass and isotype (Total IgG, IgA1 and IgA2; Southern Biotech). After 1 hour incubation, plates were washed, and flow cytometry was performed with an IQue (Intellicyt), and analysis was performed on IntelliCyt ForeCyt (v8.1). The fold-change of PE median fluorescent intensity (MFI) was reported as a readout for antigen-specific antibody titers.

### Antibody-dependent neutrophil phagocytosis (ADNP)

*S*. Enteritidis FliC was biotinylated using Sulfo-NHS-LC-LC biotin (Thermo Fisher) and coupled to fluorescent Neutravidin-conjugated beads (Thermo Fisher). To form immune complexes, antigen-coupled beads were incubated for 2 hours at 37°C with diluted serum samples (1:25) and then washed to remove unbound immunoglobulins. Immune complexes were incubated for 1 hour with RBC-lyzed whole blood at 37°C. Following the incubation, RBC-lyzed whole blood was washed, stained for CD66b (Biolegend) to identify neutrophils, and then fixed in 4% PFA. Flow cytometry was performed to identify the percentage of cells that had phagocytosed beads as well as the number of beads that had been phagocytosis (phagocytosis score = % positive cells × Median Fluorescent Intensity of positive cells/10000). Flow cytometry was performed with an IQue (Intellicyt), and analysis was performed on IntelliCyt ForeCyt (v8.1) or using FlowJo V10.7.1.

### Statistical Analysis

No formal power calculations or hypothesis testing was performed. The sample size was deemed appropriate to meet the objectives of this first-in-human study. The safety analysis was performed on all participants that received blinded study product. The immunogenicity analysis was performed on all participants that contributed blood specimen at any point post-vaccination. Other than phone call visits performed in lieu of in-person visits due to pandemic restrictions, there were no major protocol deviations.

Demographics and baseline characteristics were summarized using descriptive statistics and safety and immunogenicity by summary statistics.

## Results

### Trial Population

Between 09 December 2019 and 20 February 2020, 22 participants were enrolled and randomly allocated to blinded product groups (CONSORT diagram **Supplementary Fig. 1**). Eight participants received 6.25 μg TSCV and 2 received placebo in Cohort A (8 females and 2 males; mean age 31.6 years, range 22-44 years; 60% African American, 20% White, 10% Asian, none were Hispanic) and 10 participants received 12.5 μg TSCV and 2 received placebo in Cohort B (9 females and 3 males; mean age 31.6 years, range 18-45 years; 50% African American, 42% White, 8% declined race, none were Hispanic) (**Supplementary Table 1**). During the follow-up period, SARS-CoV-2 pandemic restrictions precluded the ability to complete planned in-clinic visits, so safety assessments were conducted by telephone. All 22 participants are included in the safety analysis. Only 20 participants are included in the primary immunogenicity analysis because 2 individuals in the 12.5 µg TSCV group could not provide a blood specimen at the scheduled Day 29 visit. Once pandemic restrictions were relaxed, 18 participants consented for a single supplemental in-clinic visit between 28 April 2021 and 07 May 2021, facilitating the assessment of persistence of immune responses to the single vaccination.

### Safety

No serious adverse events (SAEs) were reported during the study, including at supplemental visits which occurred >1 year from vaccination. All solicited reactions were mild or moderate; none of the reactions extended beyond 7 days post-vaccination (**Supplementary Table 2, Supplementary Figures 2 and 3**). Pain (59%) and erythema (41%) were the most common local injection-site reactions. Fatigue (27%), myalgia (27%), headache (27%), and malaise (18%) were most common systemic reactions; no fevers (oral temperature >100°F) were recorded. Within Cohort B, 3 participants experienced moderate myalgia within 3 days of vaccination. These 3 events took place in late February 2020, were brief, and self-resolved within 24 hours; none were associated with fever or laboratory abnormalities. There were 3 non-solicited, non-serious adverse events (AEs) experienced by 2 participants in Cohort A and 6 non-solicited AEs among 4 participants in Cohort B. The one AE considered definitively related to vaccination was the finding of mild subcutaneous swelling which was located anterior to the injection site and thus not classified as injection site induration. There were 3 AEs considered probably related to vaccination; 2 AEs were moderate-grade abnormal C-Reactive Protein (CRP) values (moderate grade CRP was defined as between 2-5 times the upper limit of the normal range; the lab reference normal was <0.8 mg/dL), and 1 AE report described mild “heaviness in the arm” (arm of vaccine receipt) the evening of vaccination. There were no trends indicating dose-dependent safety concerns.

Among participants receiving 6.25 μg TSCV, 1 participant had a CRP of 2.3 mg/dL 1-day post-vaccination which at 7-days post-vaccination was 0.5 mg/dL; this was the same person experiencing the mild subcutaneous swelling AE of the arm. Among participants receiving 12.5 μg TSCV, 4 had elevated CRP 1-day post-vaccination, range 1.2 to 2.7 mg/dL. One of these participants had CRPs of 1.0-1.1 mg/dL at screening and baseline (pre-vaccination), then CRP of 2.7 and 1.5 mg/dL at 1-day and 7-days post-vaccination. None of these 4 participants with CRP elevations were associated with other symptoms. There were no other notable clinical safety laboratory abnormalities detected. (**Supplementary Figure 4**)

### Antigen-specific serum IgG enzyme-linked immunosorbent assay (ELISA) responses

The primary immunological endpoint was seroconversion (defined as four-fold or higher titers above baseline) in serum IgG antibodies to *S*. Enteritidis (SE) and *S*. Typhimurium (STm) COPS and *S*. Typhi Vi (primary antigens) and to SE and STm FliC (secondary antigens) measured by ELISA, at 28-days post-vaccination (Day 29). In response to the 6.25 μg TSCV, there were 100% (8/8) seroconversions in IgG specific to all 3 primary antigens; 87.5% (7/8) seroconversions in both anti-SE and -STm FliC IgG. Following the receipt of 12.5 μg TSCV doses, there were 100% (8/8) seroconversions in IgG specific to the 3 primary and two secondary antigens. None of the 4 placebo recipients (0/4, 0%) exhibited IgG seroconversions to any primary or secondary antigen. [**Figure 1A**]

**Figure 1.**
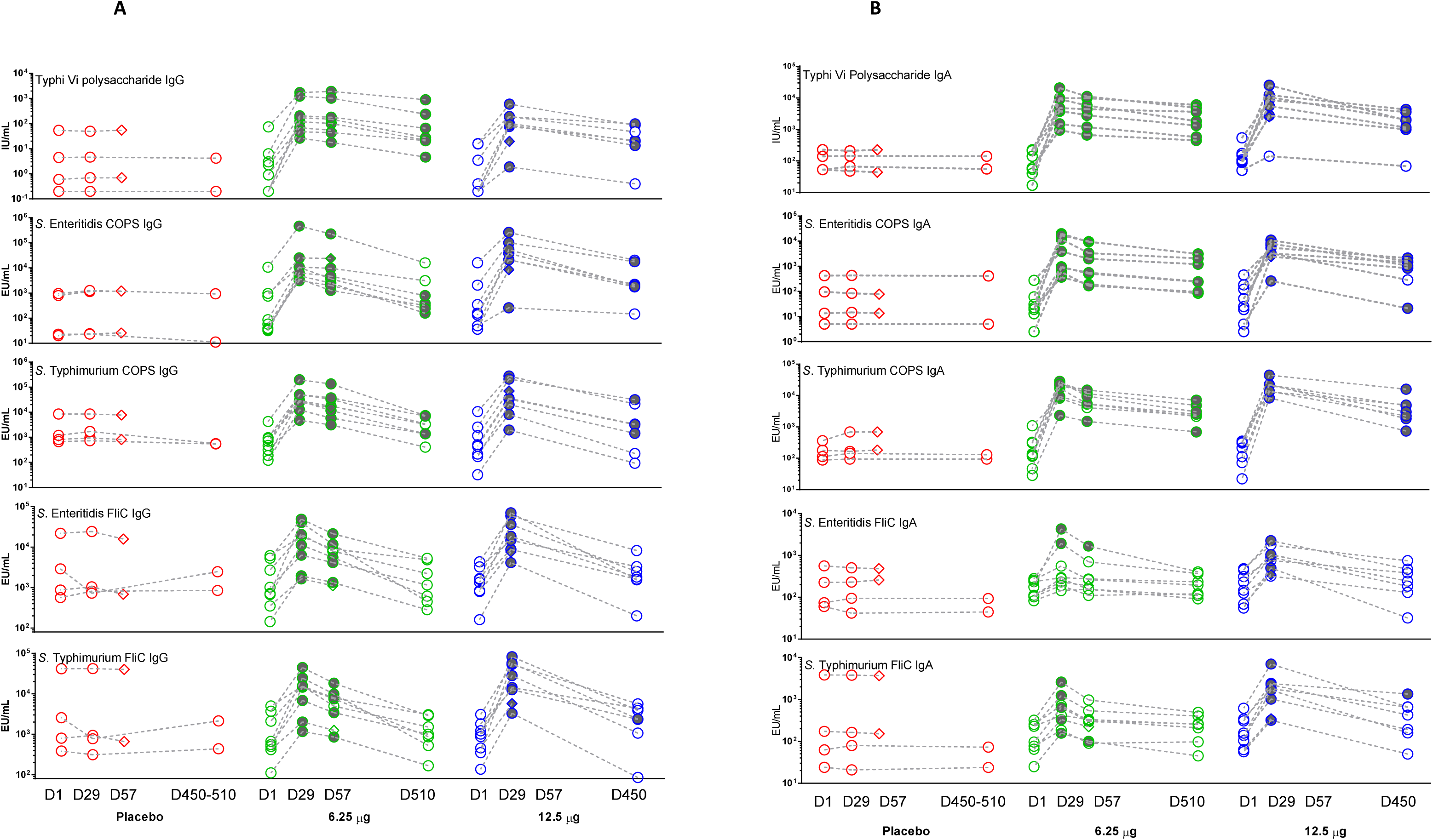
Serum antigen-specific ELISA antibody responses. Individual serum antigen-specific IgG (**Panel A**) and IgA (**Panel B**) titers were measured by ELISA, expressed as ELISA units per mL (EU/mL) or international units per ml (IU/mL), subsequent to a single dose of placebo or 6.25 μg or 12.5 μg of trivalent *Salmonella* conjugate vaccine (TSCV) at 4 timepoints: Day 1 (pre-vaccination), 29, 57, and 450-510. Shaded shapes indicate timepoints where there was a seroconversion (four-fold or higher titer than baseline titer). Diamond shapes indicate subjects missing the day 450-510 timepoint.

At 56-days post-vaccination (Day 57), the antigen-specific IgG responses were measured for the 6.25 μg TSCV recipients but were unable to be determined for the 12.5 μg TSCV recipients (Cohort B) because no serum was collected at this timepoint, due to pandemic closures. The seroconversion rates of recipients of 6.25 μg TSCV were 100% (8/8) for all 3 primary antigens and 62.5% (5/8) for the 2 secondary antigens. Placebo recipients demonstrated no seroconversions (0/2) in IgG specific to all 3 primary and the 2 secondary antigens.

Because of the prolonged pandemic closures, no specimens could be collected at a planned 180- days post-vaccination visit. However, following the rescinding of pandemic restrictions, we could collect serum >1-year post-vaccination (at ∼450 days and ∼510 days respectively for the 2 cohorts).

Serendipitously, we were able to document persistence of antigen-specific IgG antibody elevations, as titers that remained above their baseline (pre-vaccination) levels. The persistence of antibody ≥4-fold over baseline at ∼510 days post-vaccination among recipients of 6.25 μg TSCV was 71.4% (5/7) for anti-SE COPS, 42.9% (3/7) for anti-STm COPS, and 100% (5/7) for anti-Vi; however, 0% (0/7) vaccinees had elevated IgG titers against the 2 secondary antigens. The proportion of subjects vaccinated with 12.5 μg of TSCV whose titers remained ≥4-fold over baseline at ∼450-days post-vaccination was 71.4% (5/7) for anti-SE COPS, 57.1% (4/7) for anti-STm COPS, and 71.4% (5/7) for anti-Vi but was 0% (0/7) for anti-SE FliC and only 14.3% (1/7) for anti-STm FliC. Among the placebo recipients none had IgG titers above baseline (0/2) specific to any of the 5 antigens. [**Figure 1A** and **Supplementary Table 3 and Supplementary Figure 5**]

### Antigen-specific serum IgA ELISA responses

The measurement of IgA response was not a pre-defined primary immunological endpoint, as the hypothesis was that predominantly IgG responses would be elicited from this parenteral vaccine. However, at 28 days post-vaccination with 6.25 μg of TSCV, 100% (8/8) of subjects exhibited seroconversions in their titer of serum IgA against all 3 primary antigens; 25% (2/8) seroconverted for IgA anti-SE FliC and 62.5% (5/8) for IgA anti-STm FliC. Among participants who received 12.5 μg of TSCV, 100% manifested IgA seroconversion at Day 28 for both anti-SE and -STm COPS, 87.5% (7/8) for anti-Vi, 75% (6/8) for anti-SE FliC and 100% (8/8) for anti-STm FliC IgA responses. There were no seroconversions (0/4) of serum IgA specific to any of the 3 primary or 2 secondary antigens among the placebo recipients.

At 56 days post-vaccination (Day 57) the proportion of subjects with persisting IgA titers ≥4-fold above baseline among recipients of 6.25 μg TSCV was 100% (8/8) for all 3 primary antigens and 12.5% (1/8) for both secondary antigens. No placebo recipients exhibited IgA titers ≥4-fold above baseline (0/2) on day 56 against any of the 5 antigens.

For most vaccinees the IgA anti-COPS and anti-Vi remained elevated ≥4-fold above baseline at >1-year post-vaccination. At ∼510 days post-vaccination, among recipients of 6.25 μg TSCV, IgA titers were ≥4-fold above baseline among 85.7% (6/7) for anti-SE and anti-STm COPS, 100% (7/7) for anti-Vi, and 0% (0/7) for IgA against FliC antigens. The seroconversion rate at ∼450-days post-vaccination for recipients of 12.5 μg TSCV were 85.7% (6/7) for anti-SE COPS, 100% (7/7) for anti-STm COPS, 85.7% (6/7) for anti-Vi, 0% (0/7) for anti-SE FliC, and 14.3% (1/7) for anti-STm FliC IgA responses. There were no seroconversions (0/2) in IgA specific to any of the 5 antigens among the placebo recipients. [**Figure 1B** and **Supplementary Table 3 and Supplementary Figure 6**]

### Antigen-specific antibody secreting cells (ASC) responses

Induction of ASC specific to the 3 primary (Vi, SE LPS and STm LPS) and 3 secondary antigens (SE FliC, STm FliC and TT) were measured (see Methods) in unsorted fresh PBMC collected from participants who received 12. 5 µg of TSCV (n=10) before (D1) and 7 days (D8) after immunization [**Figure 2** and **Supplementary Table 4**]. The magnitude of IgA ASC responses against Vi were higher (p<0.05) than the corresponding IgG ASC, while similar IgG and IgA ASC responses were elicited against SE LPS (p=0.38) and STm LPS (p=0.25). ASC responses to STm FliC (p=0.04) and TT (p<0.01) were mostly observed for IgG [**Figure 2A** and **Supplementary Table 4**] compared to the corresponding IgA responses [**Figure 2B** and **Supplementary Table 4**]. The responders, showing post-vaccination increases >10 spot forming cells (SFC)/million PBMC, for IgA or IgG ASC, ranged between 60-100% of vaccinees, depending on the antigen. The only exception was against SE FliC which showed only 20%-30% responders. No specific IgG or IgA ASC responses to any antigen were observed at either D1 or D8 among Placebo recipients (n=2).

**Figure 2.**
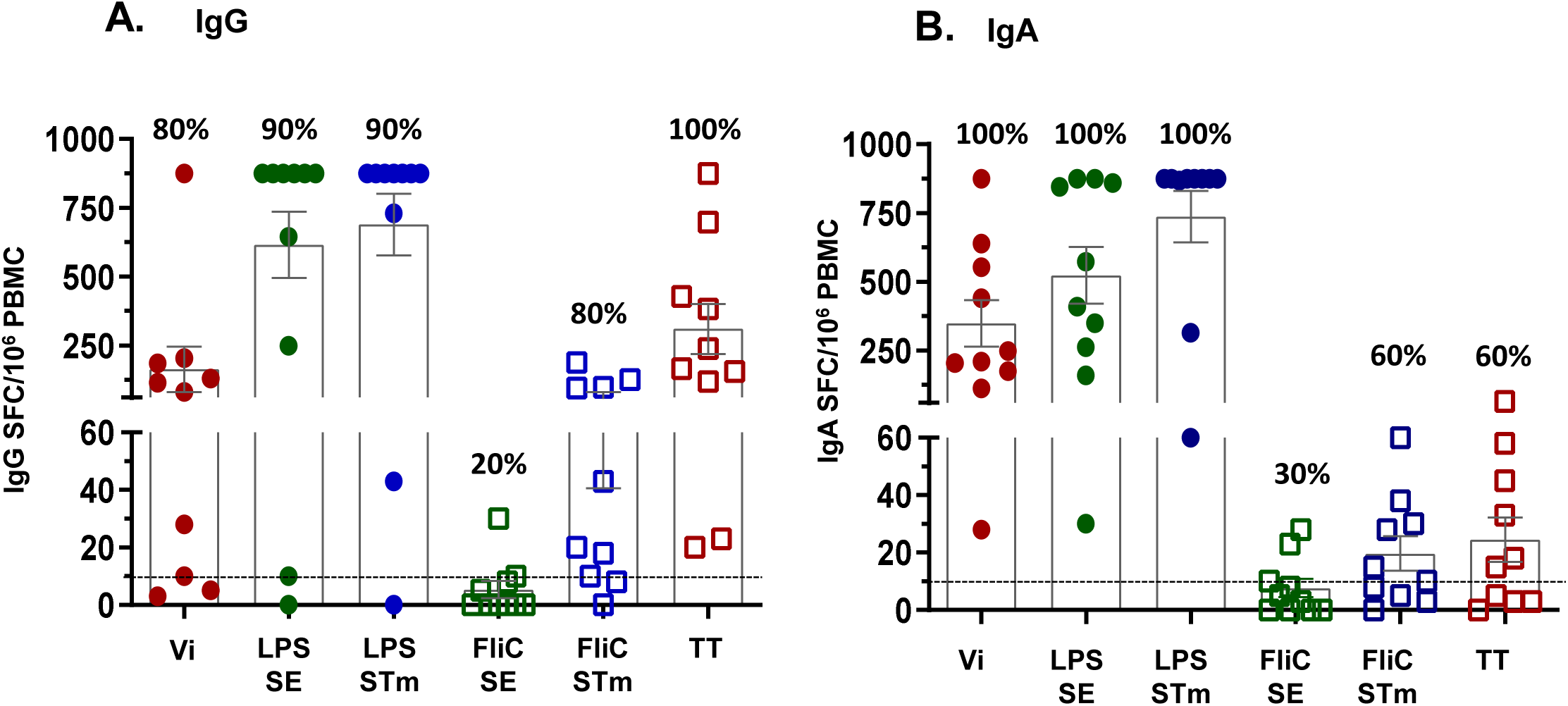
Induction of antigen-specific Antibody Secreting Cells (ASCs). ASCs were measured before (Day 1) and 7 days post-vaccination (Day 8) with 12.5 µg of TSCV intramuscularly (n=10). Data shown are net post-vaccination increases (Day 8 minus Day 1) ASC responses, defined as number of Spot Forming Cells (SFC)/10^6^ Peripheral Blood Mononuclear Cells (PBMC), for antigen-specific IgG (panel A) or IgA (panel B) ASCs. The box and error bars represent the group Mean± SEM. The horizontal dashed line represents the cutoff for positive responses (defined as ≥10 SFC/10^6^ PBMC. The percentages at the top of each bar represent the proportion of positive responders among the vaccinees (n=10). Vi: *S*. Typhi Vi Polysaccharide, LPS: Lipopolysaccharide, SE: *S*. Enteritidis, STm: *S*. Typhimurium, TT: Tetanus Toxoid.

To evaluate the tissue homing potential of TSCV-elicited antigen-specific ASC, an optimized flow cytometry-based sorting protocol was used to characterize five subpopulations of highly purified B cells, using fresh PBMC collected on post-vaccination D8 [**Supplementary Figure 7**]. The naïve B cell subset (CD19+CD27-) constituted the largest component of B cells but exhibited virtually no (0-35 SFC/million CD19+CD27- B cells) antigen-specific ASC. Whereas, antigen-specific ASC responses were observed in the 4 subsets of CD19+CD27+ B cells, showing the potential for homing to (i) lymph nodes only (CD19+CD27+CD62L+integrin α4β7-; LN), (ii) gut mucosa only (CD19+CD27+CD62L-integrin α4β7+; Gut), (iii) both lymph nodes and gut mucosa (CD19+CD27+CD62L+integrin α4β7+; LN & Gut) or (iv) other tissues (CD19+CD27+CD62L-integrin-α4β7-; undefined homing sites). [**Figure 3** and **Supplementary Table 5**] Vi-specific IgA responses observed in LN & Gut (CD62L+integrin α4β7+; p=0.02) and Gut only (CD62L-integrin α4β7+) homing subsets were both significantly (p=0.02) higher than those in the corresponding Vi-specific IgG ASC. SE LPS-specific IgA (p=0.03) and STm LPS-specific IgA (p=0.03) responses in Gut only homing subsets were significantly higher than the corresponding IgG subsets. In the LN & Gut homing subset, a similar trend for somewhat higher responses was also observed with SE LPS-specific (p=0.19) and STm LPS-specific IgA compared to IgG responses(p=0.08). Comparable magnitudes (p>0.2) of IgA and IgG ASC responses specific to Vi, SE LPS and STm LPS, were observed in the other two subsets, i.e., LN (CD62L+integrin α4β7-) and undefined homing (CD62L-integrin α4β7-) subsets.

**Figure 3.**
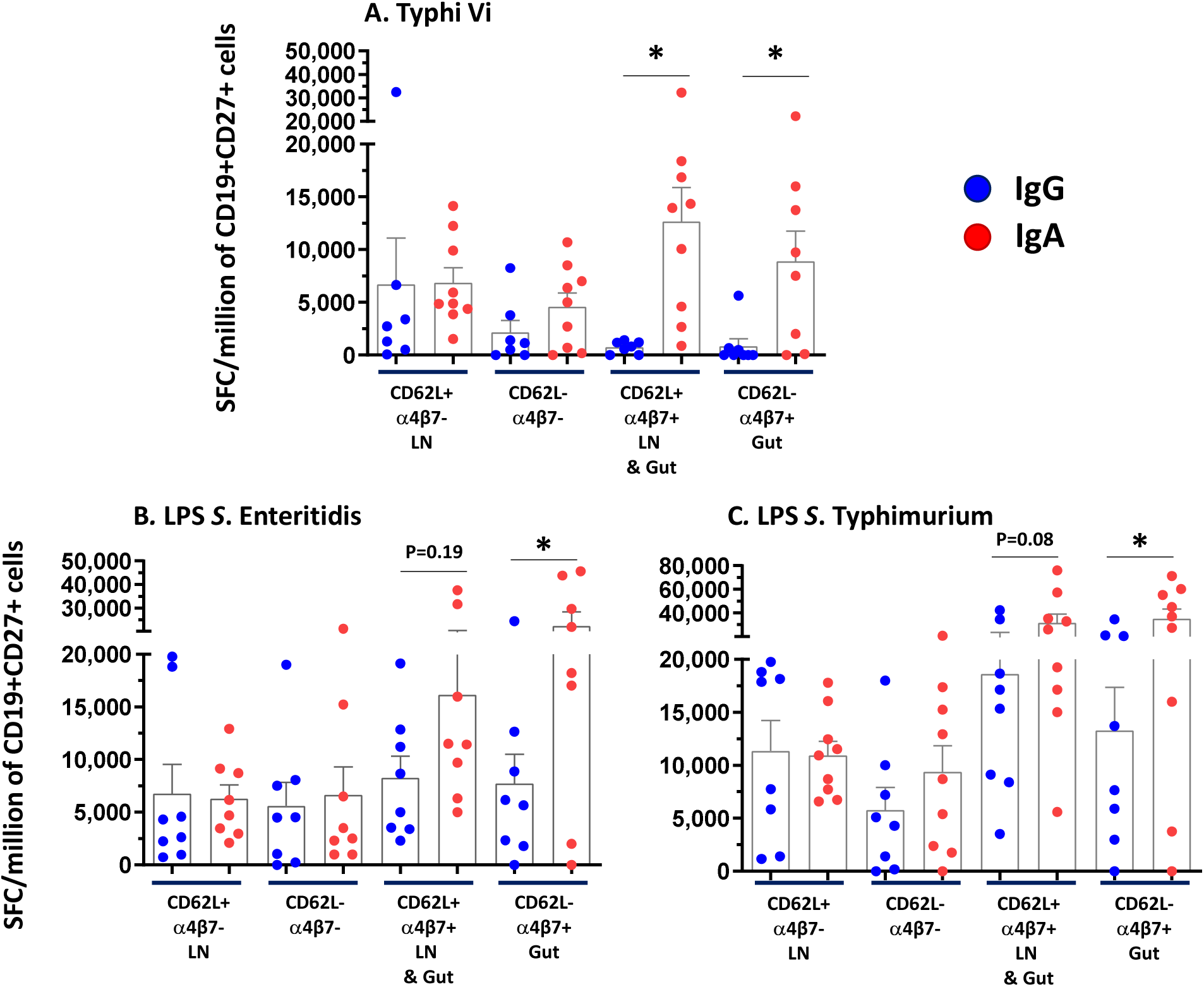
Homing of antigen-specific Antibody Secreting Cells (ASCs). On Day 8, memory B cells (CD19+ CD27+) were sorted into 4 subpopulations: CD62L+ α4β7- (LN: Lymph Node homing), CD62L- α4β7- (undefined lymphoid tissue homing), CD62L+ α4β7+ (LN and gut mucosa homing), and CD62L- α4β7+ (gut mucosa homing) from recipient of 12.5 µg of TSCV intramuscularly (n=10). Antigen-specific cells detected for each sorted subpopulation were converted to Spot Forming Cells (SFC)/million of purified CD19+ CD27+ cells based on the number of sorted cells seeded in each well. Subjects with >500 SFC/million cells in at least in 2 out of 4 sorted subsets for each antigen were included in the analysis. P values were calculated by comparing the corresponding IgG vs IgA by Wilcoxon paired test, 2-way. *p<0.05.

### B Memory (BM) Responses

The induction and persistence of polysaccharide antigen (i.e., Vi, SE LPS and STm LPS)-specific classical BM cells was evaluated from cryopreserved PBMC collected before vaccination (D1) and post-vaccination (D29, D57, and D450-510). Antigen-specific and non-specific (total) IgG and IgA secreting BM cells were measured in each sample following in vitro expansion of PBMCs and the BM frequency reported as the percentage of corresponding total IgG or IgA producing cells (see Methods). In general, induction of IgG or IgA BM responses to the three polysaccharide antigens observed in D29 samples (from both dosage cohorts), were elevated, either significantly (p<0.05, **Figure 4** and **Supplementary Table 6**) or with a positive trend (p<0.1, **Figure 4, panel E**), compared to the corresponding pre-vaccination (D1) levels. The only exception being STm LPS IgG responses among the 12.5 µg TSCV recipients (**Figure 4, panel F**). The persistence of BM cells on D57, was only observed from the 6.25 µg TSCV recipients (as no PBMC were collected for this time point among the 12.5 µg TSCV cohort due to COVID-19 pandemic restrictions). Despite a decline in the magnitude of BM responses from the peak at D29, Vi (p<0.05, **Figure 4, panels A and G**), SE LPS (p<0.05, **Figure 4, Panel H**), and STm LPS (p=0.06, **Figure 4, Panels C and I**) specific BM responses observed at D57 remained higher than the corresponding D1 levels. BM responses among most vaccinees declined to pre-vaccination levels by D450/510. The percentages of IgG and IgA BM responders (showing a post-vaccination increase in at least one post-vaccination timepoint) varied according to the antigen evaluated. The percentages of responders among the 6.25 µg TSCV recipients for Vi- (**Supplementary Figure 8, Panel A**), SE LPS- (**Supplementary Figure 8, Panel B**) and STm LPS-specific (**Supplementary Figure 8, Panel C**) IgG or IgA BM were significantly (all p<0.05) higher than in placebo recipients. Similar trends were also observed in the 12.5 µg group. We observed virtually no BM responses against LPS from *S*. Choleraesuis, which was a negative control for the BM assay, among vaccinees or placebo recipients.

**Figure 4.**
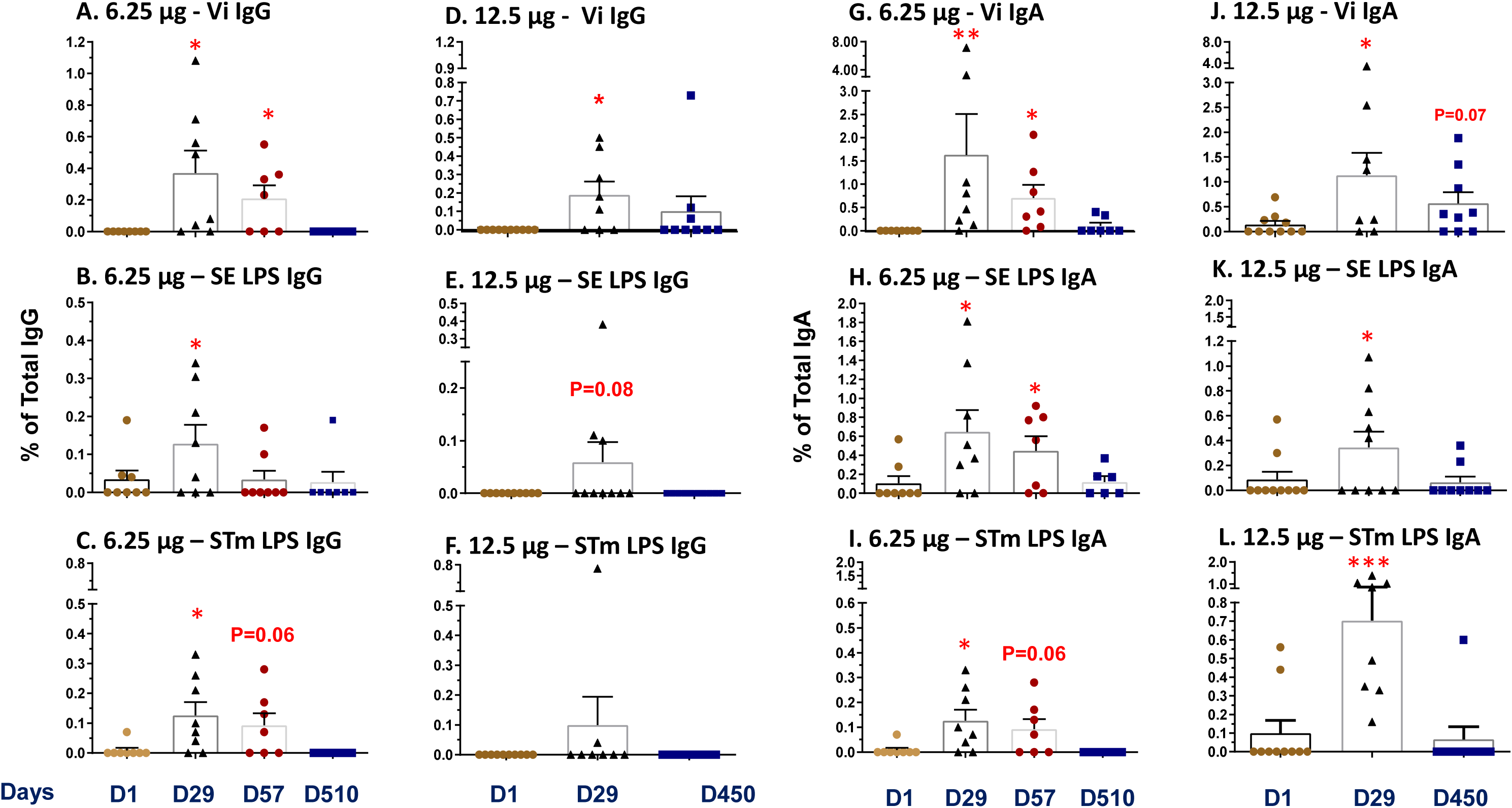
Induction of B memory (BM) cells. Antigen-specific IgG or IgA expressing BM cells were measured before (Day 1) and on Days 29, 57, or greater than 1 year (Days 450-510) after vaccination against the 3 primary antigens: Typhi Vi polysaccharide, *S*. Enteritidis LPS, or *S*. Typhimurium LPS. Antigen-specific BM cells are presented as the percentage of their corresponding total IgG or IgA levels (see Methods). Post-vaccination values were compared to the corresponding pre-vaccination values by paired t tests. *p<0.05, **p<0.01, ***p<0.001. Bars show the mean and standard error for each group and time point.

### Antibody Profiling

To further discriminate some characteristics of the antibody responses to TSCV, a Luminex-based assay was utilized. Among vaccinees, antigen-specific total IgG responses were demonstrated by all vaccine antigens and the positive control (*Citrobacter freundii* Vi polysaccharide which mimics Typhi Vi polysaccharide), while there was a general lack of responses among vaccinees to the negative control antigen (*S*. Choleraesuis LPS) and among placebo recipients to any antigen. [**Figure 5**] However, there was a single vaccinee that demonstrated a rise with the negative control antigen, which could be due to cross reactive antibody to an irrelevant gut colonizing *Enterobacteriaceae* with an LPS structure which mimics *S*. Choleraesuis,^18,19^ as no alternate explanation was discovered. These data were consistent with the ELISA antibody responses recorded, providing confidence of the Luminex assay’s readout.

**Figure 5.**
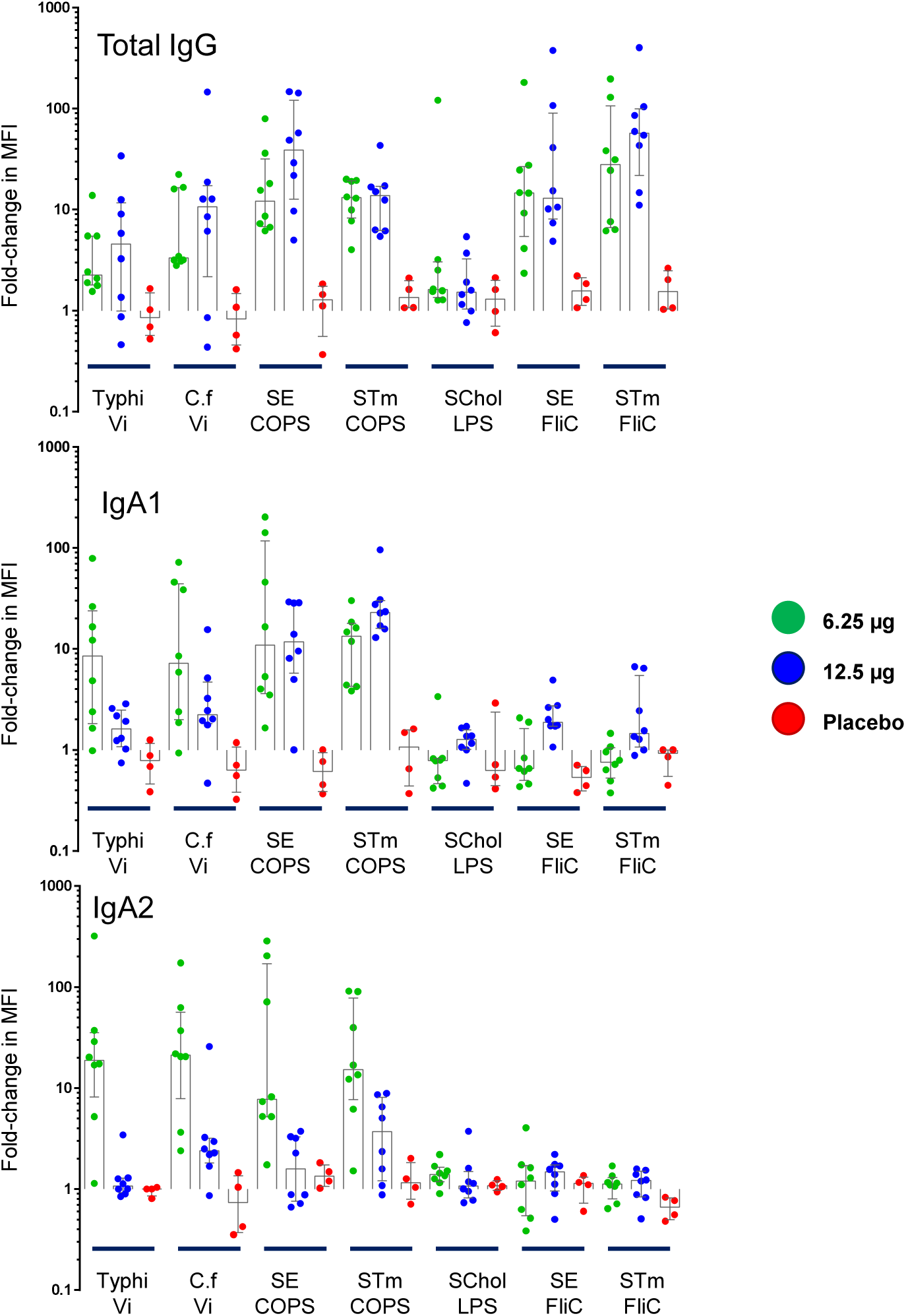
Luminex Antibody Profiling. Antibody responses were assessed with a Luminex-based assay for antigen-specific total IgG (**Panel A**) and IgA1 and IgA2 subclass (**Panel B** and **Panel C**). Data are expressed as the fold-change in median fluorescent intensity (MFI) between the baseline (Day 1) and Day 29 serum samples. Vi: Vi Polysaccharide, C.f: *Citrobacter freundii* Vi Polysaccharide, SE: *S*. Enteritidis, STm: *S*. Typhimurium, SChol: *S*. Choleraesuis, COPS: Core and O-Polysaccharide, LPS: Lipopolysaccharide.

The antigen-specific IgA subclasses were also evaluated. Vaccinees showed balanced responses between the polysaccharide-specific IgA1 and IgA2 levels elicited. [**Figure 5**] However, there were weak responses against the FliC components, and they were predominantly IgA1 subclass; only a single vaccinee demonstrated an SE FliC IgA2 response. Placebo recipients lacked IgA subclass responses to any antigen.

The functional capacity for antibody-dependent neutrophil phagocytosis (ADNP) was only assessed with respect to SE FliC [**Figure 6**]. There was evidence for ADNP responses elicited 1-month post-vaccination. However, these antibodies decreased at 2 months post-vaccination and virtually disappeared after 1 year. The safety, clinical acceptability and striking immunogenicity of single fractional doses of TSCV in the aborted Phase 1 trial nevertheless paved the way for a Phase 2 trial in North American adults that included a Half-strength GMP formulation of TSCV, as well as a new Full-strength GMP formulation, and a bridging group of subjects who received dilutional Half-strength as prepared in the Phase 1 study.

**Figure 6.**
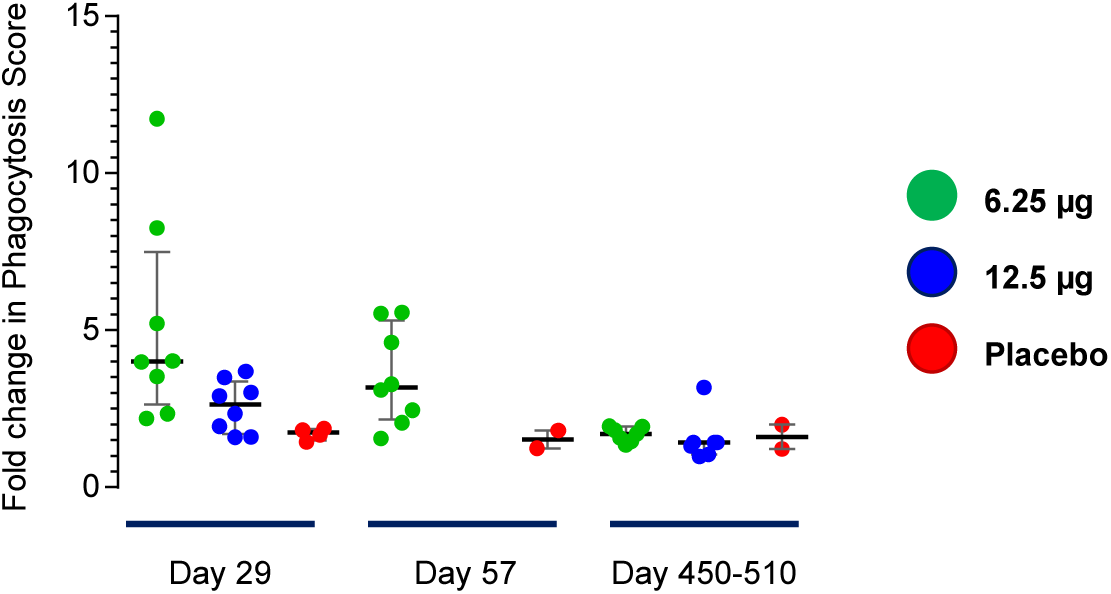
Antibody Dependent Neutrophil Phagocytosis Responses. Immune complexes, consisting of serum mixed with biotinylated *S*. Enteritidis FliC and tagged Neutravidin-conjugated beads, were incubated with neutrophils. Then, flow cytometry was performed to measure the percentage of phagocytosed beads. Data are expressed as the fold-change in phagocytosis score = % positive cells × Median Fluorescent Intensity of positive cells/10,000 between the baseline (Day 1) and Day 29 serum samples.

## Discussion

Recognition during the past three decades of the frequency and severity of iNTS disease among infants and toddlers in sub-Saharan Africa,^20,21^ revelation of distinct genomic features of African iNTS versus NTS from North America and Europe,^22,23^ and elucidation that the reservoir of African iNTS is apparently restricted to human hosts (like typhoidal serovars)^24^ has stimulated efforts to develop vaccines to prevent iNTS disease. TSCV combines conjugates that utilize COPS antigens from serogroup O:4 serovar *S*. Typhimurium and from serogroup O:9 serovar *S.* Enteritidis to elicit O antibody responses to these most prevalent iNTS serovars. However, these O antigens are also expected to protect against other distinct serovars within serogroup O:4 (e.g., I:4,5:i:- and I:4,5:nm) and against distinct serovars within serogroup O:9 that are prevalent (e.g., *S*. Dublin).

Innovations of the CVD-Bharat TSCV include the use of serovar-homologous FliC as the carrier protein to strengthen protection and use of reagent strains to simplify and economize manufacture of the iNTS conjugates. Combination of the two iNTS conjugates with licensed Typbar TCV (Vi-TT) achieves a trivalent conjugate that intended, collectively, to elicit protection against all the major invasive *Salmonella* pathogens that threaten young children in sub-Saharan Africa. Herein we showed the results from the first-in-human Phase 1 clinical trial in North American adults.

The Phase 1 trial tested only quarter-strength and half-strength dose levels of TSCV because of extended COVID-19 prohibitions that led to a university-wide lockdown and prohibition of clinical research that was not COVID-19 vaccine or treatment-related. Consequently, the full-strength dose level (25 µg per conjugate) was not evaluated in this study. These COVID-19-related interruptions led to delays in collection of closeout blood specimens far beyond the Day 180 per protocol date.

Consequently, not only was safety documented and Day 29 seroconversions recorded but, serendipitously, when clinical visits were finally allowed on ∼Day 510 (Cohort A) and ∼Day 450 (Cohort B) to collect sera, long-term elevations of specific anti-COPS and anti-Vi antibody were observed after a single quarter-strength and a half-strength dose level of TSCV. This raised plausibility that the dose level of the two iNTS conjugates in future studies could be diminished by 50%, thereby diminishing the cost of goods and increasing the future supply of vaccine, parameters of immense importance for a vaccine targeting infants and toddlers in some of the world’s least-resourced countries in sub-Saharan Africa.

A limitation of this study is the extremely small sample sizes of the study. The original design of the protocol not only described the evaluation of the full-strength dose level, but also the evaluation of a single-dose versus a two-dose regimen spaced 28 days apart. The adaptive design also included a final “confirmatory” cohort which would have significantly increased the number of participants exposed the TSCV. Were the study uninterrupted, the original design would have enrolled a total of 104 individuals, among which 88 would have received TSCV and 16 received placebo. Therefore, the small sample size of this study restricts the ability to make definitive conclusions regarding the safety and immunogenicity of TSCV. The generalizability of the study findings will only be proven with additional clinical studies of TSCV.

The use of Phase 1 FliC as the carrier protein enhances the array of protective antibodies stimulated. However, the use of the FliC component raised a theoretical safety question since Phase 2 flagellin of *Salmonella* Typhimurium when used as an adjuvant in genetic fusions with influenza virus antigens proved to be moderately reactogenic in clinical trials in U.S. adults who exhibited marked rises in TLR5 activity accompanying the reactogenicity.^25–27^ In contrast, TSCV was clinically well tolerated at both dose levels tested. The reason is that the conjugation procedure employed in the manufacture of TSCV inactivates the TLR-activating moiety of FliC. Hence the clinical acceptability of TSCV and lack of notable TLR5 elevations in contrast to the marked reactogenicity and TLR5 elevations reported with influenza virus protein-Phase 2 flagellin genetic fusion vaccines.^25–27^

The immunogenicity of the TSCV vaccine was further evidenced by the induction of robust IgG and IgA ASC responses to the Vi polysaccharide and the LPS of both SE and STm, as well as against its protein conjugate components. Interestingly, expression of the gut homing marker integrin α4β7 and/or the lymph node homing marker CD62L by circulating ASC, especially those expressing the IgA isotype, suggests an induction of cells with the ability to home to both the gut mucosal and systemic sites in spite of being a parenteral immunization. The robust IgA response was represented by a balance of IgA subclasses elicited. The quality of the serum IgA responses may be predictive of the protection afforded by TSCV against iNTS disease, as the serum IgA responses against Vi polysaccharide correlated with protection in a typhoid challenge model.^28^ Furthermore, the detection of ADNP directed against FliC provided a potential mechanistic readout for the antibodies elicited. We are not aware that ADNP has been detected against FliC previously.

The immunogenicity of TSCV in US adults was impressive, particularly for responses to the polysaccharide antigens regarded as the primary mediators of protection. Also notable is the longevity of the anti-COPS and anti-Vi IgG responses stimulated by fractional doses of TSCV. The demonstration of α4β7 markers on ASCs indicating homing of these cells to the intestinal mucosa provides evidence for the elicitation of protection at the level of the gut as well as systemic protection mediated by serum IgG antibodies.^29–31^ The B memory cell responses are also very encouraging as they indicate the elicitation of immunologic memory at the cellular level and imply the ability of the vaccinated host to mount an anamnestic response upon exposure to *Salmonella* O:4 and O:9 pathogens.

Animal studies that show the protective capacity of serum anti-FliC antibodies elicited by TSCV^32^ engender optimism that anti-FliC may also play a protective role in humans. Our demonstration that anti-FliC antibodies in conjunction with polymorphonuclear leukocytes results in killing of iNTS *in vitro*, whereas baseline sera exhibited no protective effect provide one likely mediator of the protective effect of anti-FliC antibodies.

In conclusion, TSCV at doses up to 12.5 µg were observed to be safe and well-tolerated. A single dose of TSCV elicited immune responses which were highly encouraging as they appeared to be long-lived and home to gastrointestinal mucosa. The strength and longevity of the anti-COPS and anti-FliC immune responses observed with quarter-strength and half-strength dilutional doses of TSCV altered the clinical development plan so that a GMP lot of Half-strength TSCV was prepared and tested for safety and immunogenicity versus Full-strength TSCV in U.S. adults and showed similar high rates of seroconversion of serum IgG antibodies to COPS and Vi antibodies stimulated by Half-strength as well as Full-strength TSCV (unpublished, NCT5525546). This vaccine is planned to be further assessed in an age- descending clinical study to document safety and immunogenicity in the target population of young infants and toddlers (unpublished, NCT05784701) in sub-Saharan Africa.

## Data Availability

All data produced in the present study are available upon reasonable request to the authors

## Acknowledgements

This work was supported by Wellcome Strategic Translation Award 095967/Z/11/Z from the Wellcome Trust (London, UK). The immunology work was also supported by U.S. National Institutes of Health (NIH, Bethesda, Maryland, U.S.A.) grants R01 AI036525, U19 AI082655 and U19AI181108 (Cooperative Center for Human Immunology [CCHI]), and by National Cancer Institute Cancer Center Support Grant (CCSG) P30CA134274 to MBS. The content is solely the responsibility of the authors and does not necessarily represent the official views of the U.S. National Institutes of Health or Wellcome Trust. Generous invaluable in-kind support was also provided by Bharat Biotech International Ltd. (Hyderabad, India).

We thank Lisa Chrisley and the CVD clinical research staff for their assistance in conducting the clinical trial, Brenda Dorsey for the management of quality assurance, and Aly Kwon for regulatory support. We also recognize the guidance and oversight of members of the Safety Monitoring Committee: Joel V. Chua, Robert H. Gilman, and Mary-Claire Roghmann. Lastly, we thank the study subjects who volunteered to participate in the study and endured follow-up through an unexpected pandemic.

## Author Contributions

WHC and MML obtained funding for the study. WHC, RE, KM, KME, and MML conceptualized the study and were responsible for the methodology of the study. WHC, RSB, RRR, MFP, MSS, RW, BB, and GA were involved in the investigation, WHC, MJS, RD, RoS, and YL were responsible for data curation, and WHC and YL performed the formal analysis for the study. Project supervision was provided by WHC and MML; project administration was performed by WHC, RSB and MML. Resources were obtained by SMT, RaS, SMB, JEG, AL, MGN, and DYR. Data validation was performed by MFP, MBS, SMT, and GA, while data visualization was conducted by WHC, MJS, RW, and BB. The drafting of the first version of the manuscript was by WHC and MML; all authors performed review and editing of the final version of the manuscript.

## Inclusion & Ethics Statement

The goal of the research is to develop a vaccine which represents a burden of disease in sub-Saharan Africa and local partners (collaborators of potential sub-Saharan African sites) have been included in the discussion of the further product development of TSCV, whereby the data from this study has been shared. The research of this study was conducted with close collaboration with Bharat Biotech International Limited, the manufacturer partner that has committed to making global vaccines available at low cost. The research of this study was conducted with both local ethical and international ethical standards in mind, as the next stage would be to conduct studies of TSCV in sub-Saharan Africa, where the continuation of capacity-building for local researchers can take place.

## Data Availability Statement

This clinical trial was registered at https://www.clinicaltrials.gov before any study participant enrollment (clinical trial identifier NCT03981952). Most data supporting the findings of this study are available within the paper and its Supplementary Information. Protected health information of study participants are not included in the paper, as they are protected data under U.S. law (The Health Insurance Portability and Accountability Act of 1996). The study team is committed to sharing access to the supporting data from external researchers who provide methodologically sound scientific proposals for use of the data; written requests may be submitted to the corresponding author.

