## Supplemental Appendix for "A phase 1 randomized, placebo-controlled trial of a combination typhoid and non-typhoidal *Salmonella* polysaccharide conjugate vaccine"

### Figure 1: CONSORT Diagram

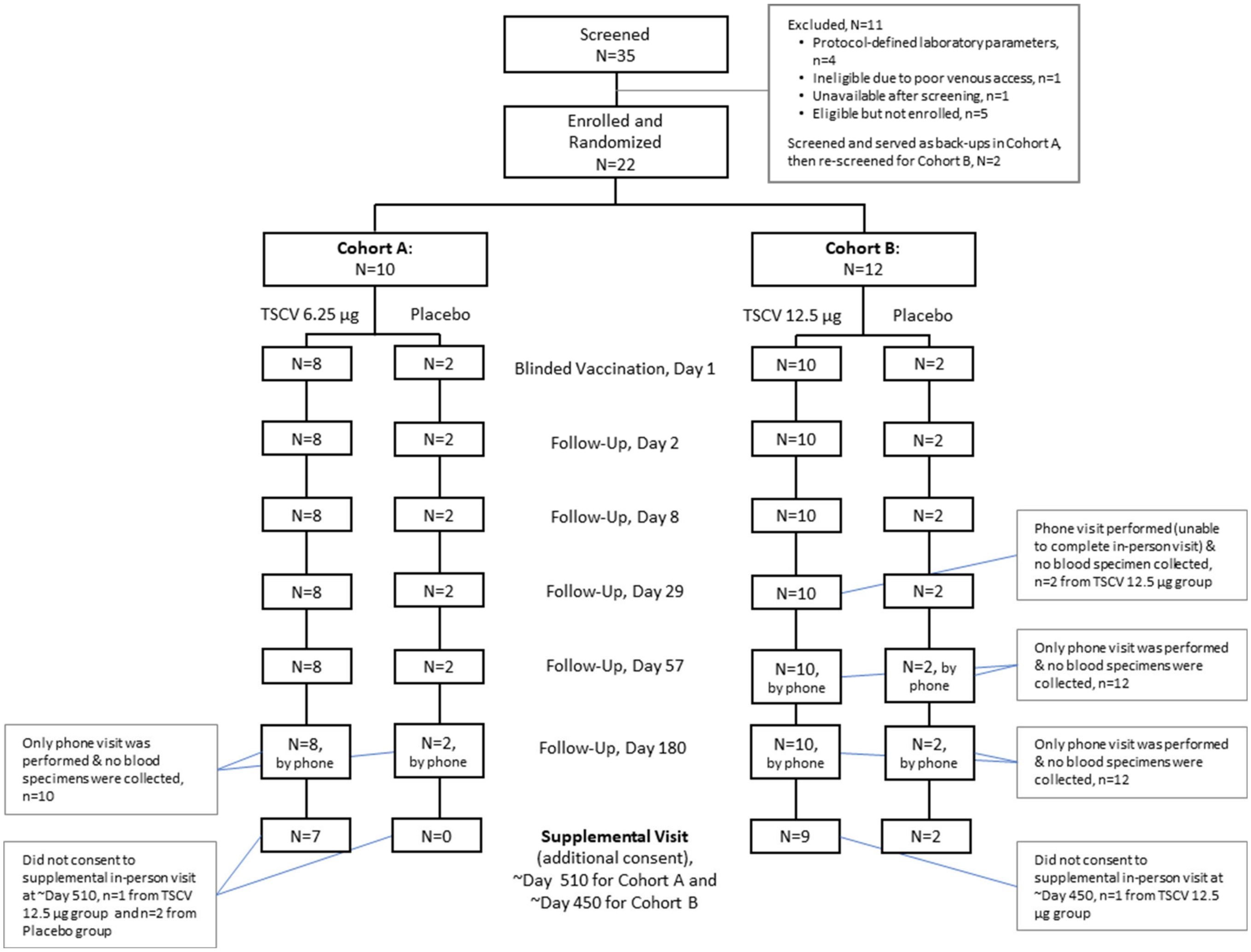

### Table 1. Baseline Demographic Characteristics

**By Cohort**

| | Cohort A TSCV 6.25 µg or placebo | Cohort B TSCV 12.5 µg or placebo | All Subjects |
|---|---|---|---|
| Total Enrolled | 10 | 12 | 22 |
| Gender, | | | |
| No. male (%) | 2 (20%) | 3 (25%) | 5 (22.7%) |
| No. female (%) | 8 (80%) | 9 (75%) | 17 (77.3%) |
| Age, year | | | |
| Mean (SD) | 31.6 (8.44) | 31.6 (9.39) | 31.6 (8.75) |
| Median [Q1, Q3] | 30.5 [23.5, 39.25] | 31 [25.25, 38.5] | 31 (23.5, 39.5) |
| Min, Max | 22, 44 | 18, 45 | 18, 45 |
| Ethnicity, | | | |
| No. non-Hispanic (%) | 10 (100%) | 12 (100%) | 22 (100%) |
| Race, n (%) | | | |
| Black/African American | 6 (60%) | 6 (50%) | 12 (54.5%) |
| White | 2 (20%) | 5 (42%) | 7 (31.8%) |
| Asian | 1 (10%) | 0 | 1 (4.5%) |
| Am Indian/Alaska Native | 0 | 0 | 0 |
| Pacific Islander/Hawaiian | 0 | 0 | 0 |
| Multi-race | 1 (10%) | 0 | 1 (4.5%) |
| Unknown or not reported | 0 | 1 (8%) | 1 (4.5%) |

**By Study Product**

| | TSCV 6.25 µg | TSCV 12.5 µg | Placebo |
|---|---|---|---|
| Total Enrolled | 8 | 10 | 4 |
| Gender, | | | |
| No. male (%) | 1 | 1 (10%) | 3 (75%) |
| No. female (%) | 7 | 9 (90%) | 1 (25%) |
| Age, year | | | |
| Mean (SD) | 33.6 (8.21) | 32.7 (9.97) | 24.7 (1.89) |
| Median [Q1, Q3] | 35 (26.75, 40.25) | 35 (24.75, 39.5) | 25.5 (24.25, 26) |
| Min, Max | 23, 44 | 18, 45 | 22, 26 |
| Ethnicity, | | | |
| No. non-Hispanic (%) | 8 (100%) | 10 (100%) | 4 (100%) |
| Race, n (%) | | | |
| Black/African American | 5 (71%) | 5 (50%) | 2 (50%) |
| White | 2 (29%) | 4 (40%) | 1 (25%) |
| Asian | 0 | 0 | 1 (25%) |
| Am Indian/Alaska Native | 0 | 0 | 0 |
| Pacific Islander/Hawaiian | 0 | 0 | 0 |
| Multi-race | 1 (14%) | 0 | 0 |
| Unknown or not reported | 0 | 1 (10%) | 0 |

### Table 2. Solicited Adverse Events

**Maximum Grade Severity over 7-days Post-Vaccination, by Solicited Symptom**

| | TSCV 6.25 µg | | | | TSCV 12.5 µg | | | | placebo | | | |
|---|---|---|---|---|---|---|---|---|---|---|---|---|
| **Grade** | **None 0** | **Mild 1** | **Mod 2** | **Sev 3** | **None 0** | **Mild 1** | **Mod 2** | **Sev 3** | **None 0** | **Mild 1** | **Mod 2** | **Sev 3** |
| Systemic Symptoms | | | | | | | | | | | | |
| Fever | 8 | | | | 10 | | | | 4 | | | |
| Chills | 8 | | | | 0 | | 1 | | 4 | | | |
| Fatigue | 8 | | | | 5 | 3 | 2 | | 2 | 2 | | |
| Malaise | 6 | 2 | | | 8 | 1 | 1 | | 4 | | | |
| Myalgia | 6 | 2 | | | 6 | 1 | 3 | | 4 | | | |
| Arthralgia | 8 | | | | 8 | 2 | | | 4 | | | |
| Nausea | 8 | | | | 8 | 2 | | | 4 | | | |
| Headache | 5 | 3 | | | 7 | 2 | 1 | | 4 | | | |
| Local Symptoms | | | | | | | | | | | | |
| Pain | 2 | 6 | | | 1 | 7 | 2 | | 4 | | | |
| Erythema | 5 | 3 | | | 5 | 4 | 1 | | 3 | 1 | | |
| Induration | 8 | | | | 7 | 2 | 1 | | 3 | 1 | | |
| Ecchymosis | 5 | 3 | | | 9 | 1 | | | 4 | | | |

### Figure 2. Local Solicited Adverse Events

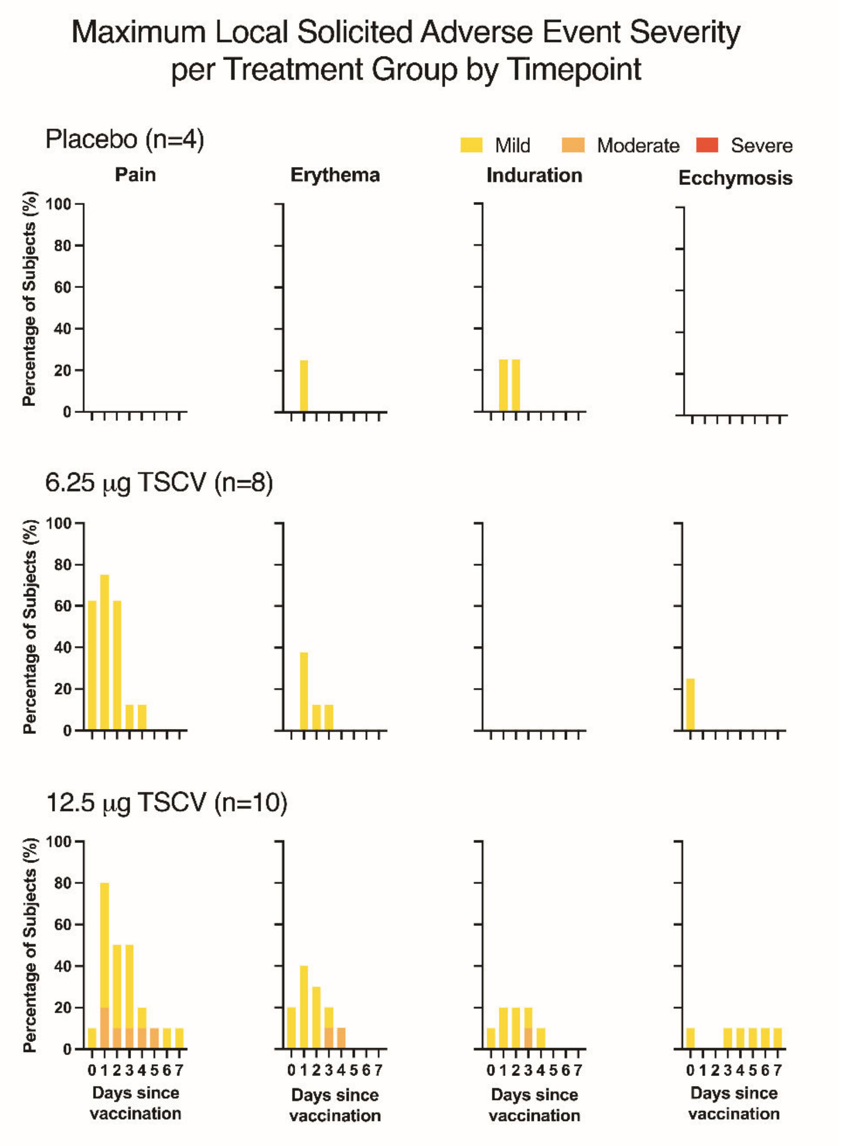

### Figure 3. Systemic Solicited Adverse Events

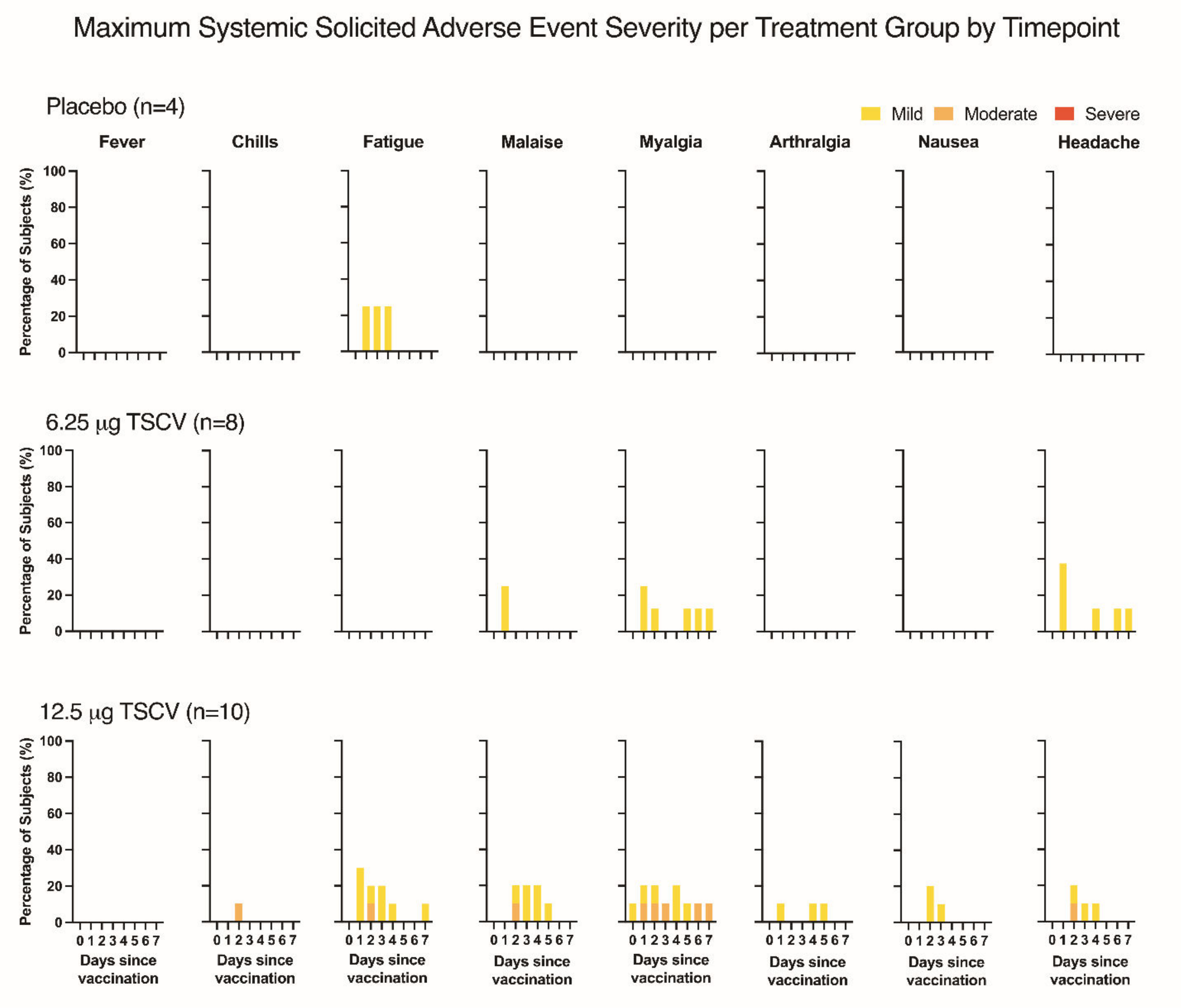

#### Figure 4. Clinical Safety Laboratories

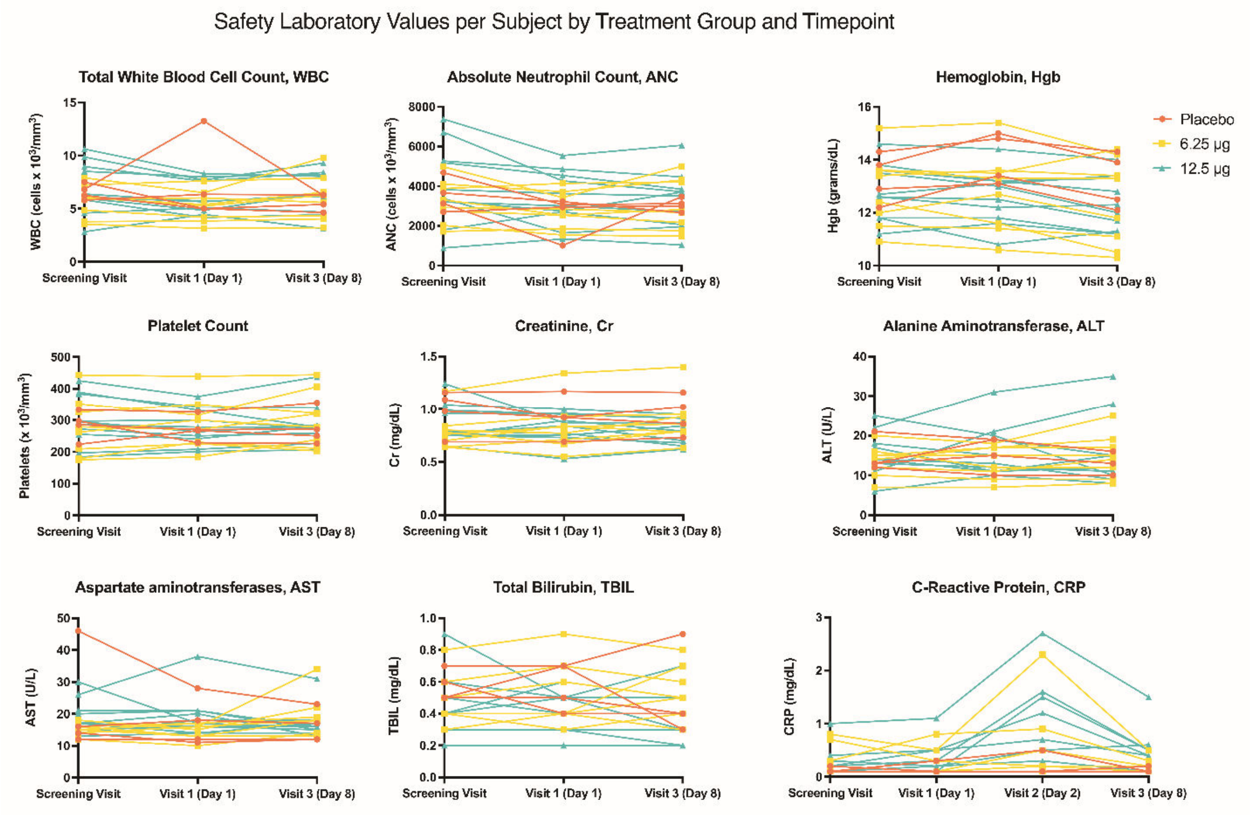

### Table 3: Serum ELISA IgG and IgA responses

***S.* Enteritidis COPS IgG (EU/mL)**

| | placebo | | 6.25 µg | | 12.5 µg | |
|---|---|---|---|---|---|---|
| | **conv** | **GMT** | **conv** | **GMT** | **conv** | **GMT** |
| Day 1 | - | 137 | - | 191 | - | 252 |
| Day 29 | 0/4 | 181 | 8/8 | 12,190 | 8/8 | 21,958 |
| Day 57 | 0/2 | 176 | 8/8 | 7,011 | ns | ns |
| Day 450/510 | 0/2 | 102 | 5/7 | 786 | 5/7 | 2,596 |

***S.* Enteritidis COPS IgA (EU/mL)**

| | placebo | | 6.25 µg | | 12.5 µg | |
|---|---|---|---|---|---|---|
| | **conv** | **GMT** | **conv** | **GMT** | **conv** | **GMT** |
| Day 1 | - | 41 | - | 25 | - | 36 |
| Day 29 | 0/4 | 40 | 8/8 | 2,709 | 8/8 | 3,501 |
| Day 57 | 0/2 | 32 | 8/8 | 1,239 | ns | ns |
| Day 450/510 | 0/2 | 45 | 6/7 | 450 | 6/7 | 594 |

***S.* Typhimurium COPS IgG (EU/mL)**

| | placebo | | 6.25 µg | | 12.5 µg | |
|---|---|---|---|---|---|---|
| | **conv** | **GMT** | **conv** | **GMT** | **conv** | **GMT** |
| Day 1 | - | 1.545 | - | 556 | - | 553 |
| Day 29 | 0/4 | 1,781 | 8/8 | 29,976 | 8/8 | 33,456 |
| Day 57 | 0/2 | 2,522 | 8/8 | 17,665 | ns | ns |
| Day 450/510 | 0/2 | 553 | 3/7 | 2,398 | 4/7 | 2,176 |

***S.* Typhimurium COPS IgA (EU/mL)**

| | placebo | | 6.25 µg | | 12.5 µg | |
|---|---|---|---|---|---|---|
| | **conv** | **GMT** | **conv** | **GMT** | **conv** | **GMT** |
| Day 1 | - | 160 | - | 153 | - | 160 |
| Day 29 | 0/4 | 199 | 8/8 | 13,592 | 8/8 | 17,874 |
| Day 57 | 0/2 | 357 | 7/8 | 6,633 | ns | ns |
| Day 450/510 | 0/2 | 111 | 6/7 | 2,647 | 7/7 | 3,205 |

***S.* Typhi Vi IgG (EU/mL)**

| | placebo | | 6.25 µg | | 12.5 µg | |
|---|---|---|---|---|---|---|
| | **conv** | **GMT** | **conv** | **GMT** | **conv** | **GMT** |
| Day 1 | - | 2.4 | - | 1.4 | - | 0.9 |
| Day 29 | 0/4 | 2.4 | 8/8 | 173 | 8/8 | 70 |
| Day 57 | 0/2 | 6.1 | 8/8 | 148 | ns | ns |
| Day 450/510 | 0/2 | 1.0 | 7/7 | 50 | 5/7 | 19 |

***S.* Typhi Vi IgA (EU/mL)**

| | placebo | | 6.25 µg | | 12.5 µg | |
|---|---|---|---|---|---|---|
| | **conv** | **GMT** | **conv** | **GMT** | **conv** | **GMT** |
| Day 1 | - | 97 | - | 82 | - | 128 |
| Day 29 | 0/4 | 99 | 8/8 | 4,573 | 7/8 | 4,470 |
| Day 57 | 0/2 | 100 | 8/8 | 3,420 | ns | ns |
| Day 450/510 | 0/2 | 88 | 7/7 | 1,830 | 6/7 | 1,257 |

***S.* Enteritidis FliC IgG (EU/mL)**

| | placebo | | 6.25 µg | | 12.5 µg | |
|---|---|---|---|---|---|---|
| | **conv** | **GMT** | **conv** | **GMT** | **conv** | **GMT** |
| Day 1 | - | 2,369 | - | 1,106 | - | 1,238 |
| Day 29 | 0/4 | 1,970 | 7/8 | 10,687 | 8/8 | 18,056 |
| Day 57 | 0/2 | 3,277 | 5/8 | 5,295 | ns | ns |
| Day 450/510 | 0/2 | 1,450 | 0/7 | 1,295 | 0/7 | 1,803 |

***S.* Enteritidis FliC IgA (EU/mL)**

| | placebo | | 6.25 µg | | 12.5 µg | |
|---|---|---|---|---|---|---|
| | **conv** | **GMT** | **conv** | **GMT** | **conv** | **GMT** |
| Day 1 | - | 155 | - | 154 | - | 18 |
| Day 29 | 0/4 | 147 | 2/8 | 472 | 6/8 | 797 |
| Day 57 | 0/2 | 356 | 1/8 | 285 | ns | ns |
| Day 450/510 | 0/2 | 65 | 0/8 | 188 | 0/7 | 221 |

***S.* Typhimurium FliC IgG (EU/mL)**

| | placebo | | 6.25 µg | | 12.5 µg | |
|---|---|---|---|---|---|---|
| | **conv** | **GMT** | **conv** | **GMT** | **conv** | **GMT** |
| Day 1 | - | 2,403 | - | 874 | - | 778 |
| Day 29 | 0/4 | 1,760 | 7/8 | 9,442 | 8/8 | 19,795 |
| Day 57 | 0/2 | 5,136 | 5/8 | 4,587 | ns | ns |
| Day 450/510 | 0/2 | 970 | 0/7 | 1,008 | 1/7 | 1,746 |

***S.* Typhimurium FliC IgA (EU/mL)**

| | placebo | | 6.25 µg | | 12.5 µg | |
|---|---|---|---|---|---|---|
| | **conv** | **GMT** | **conv** | **GMT** | **conv** | **GMT** |
| Day 1 | - | 178 | - | 122 | - | 164 |
| Day 29 | 0/4 | 179 | 5/8 | 527 | 8/8 | 1,380 |
| Day 57 | 0/2 | 746 | 1/8 | 275 | ns | ns |
| Day 450/510 | 0/2 | 42 | 0/7 | 199 | 1/7 | 325 |

conv: number with seroconversion by total number of participants at that timepoint. Seroconversion was defined as four-fold or higher titer than baseline (day 1) titer.
GMT: geometric mean titer
ns: no sample
Note: for Cohort A the last visit was ~Day 510 and for Cohort B the last visit was ~Day 450.

### Figure 5. Persistence of serum antigen-specific IgG responses

Individual serum antigen-specific IgG titers, comparing day 1 (baseline) and Day 450-510 titer. Shaded shapes indicate post-vaccination timepoints where there was seroconversion (four-fold or higher titer than baseline titer). Diamond shapes indicate subjects missing the Day 450-510 timepoint.

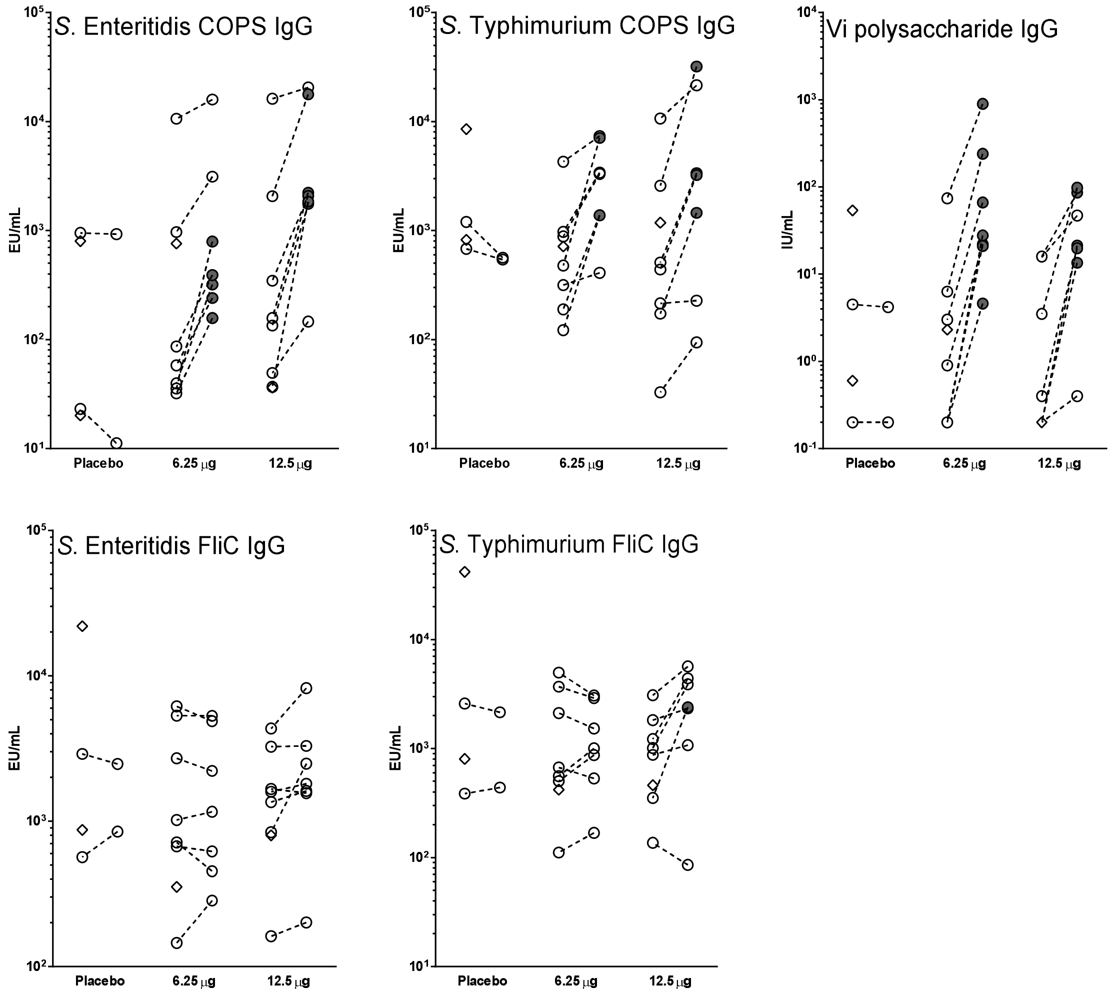

### Figure 6. Persistence of serum antigen-specific IgA responses

Individual serum antigen-specific IgA titers, comparing day 1 (baseline) and Day 450-510 titer. Shaded shapes indicate post-vaccination timepoints where there was seroconversion (four-fold or higher titer than baseline titer). Diamond shapes indicate subjects missing the Day 450-510 timepoint.

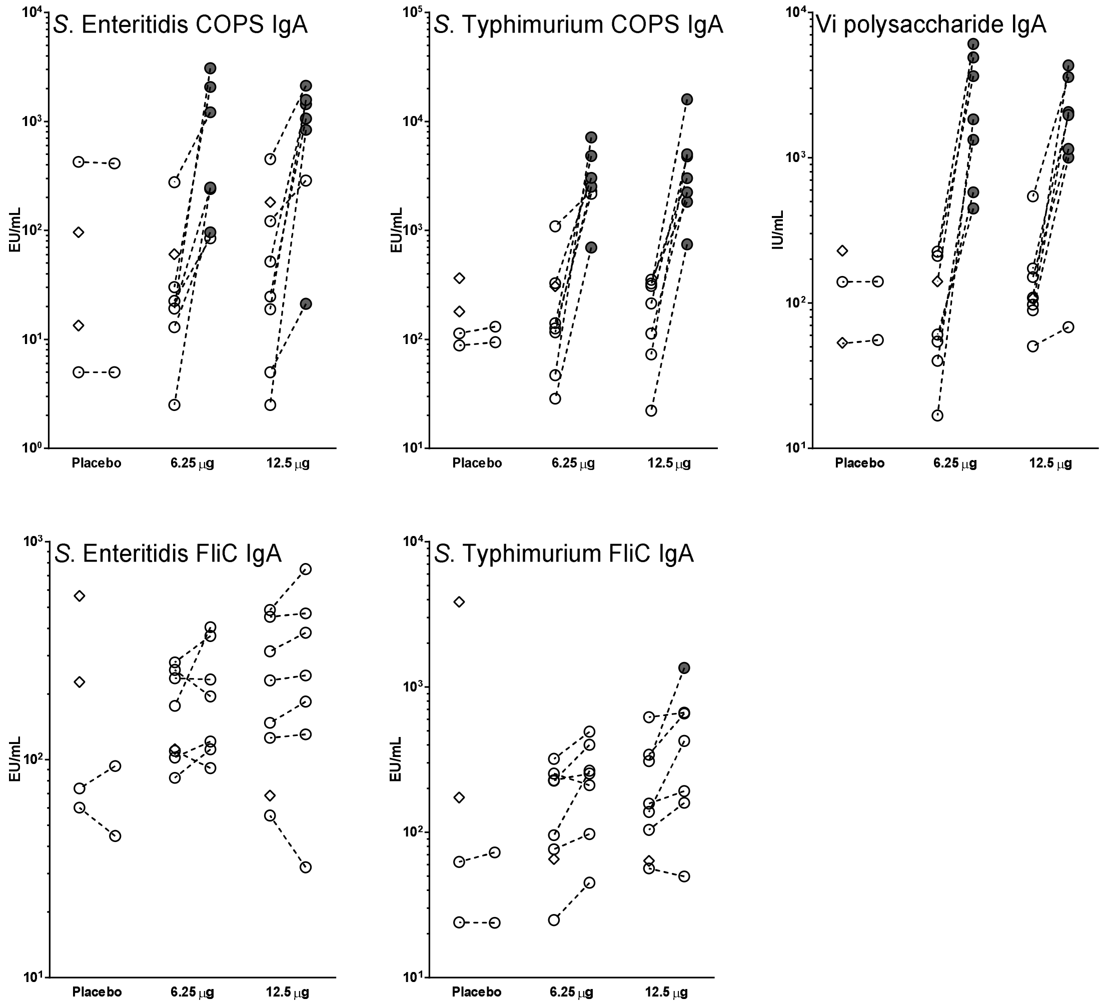

#### Table 4. Unsorted Antibody Secreting Cells Responses

Unsorted ASC responses were only measured among Cohort B participants.

12.5 µg TSCV, n=10

| ASC IgG | SE LPS | STm LPS | Vi | SE FliC | STm FliC | TT |
|---|---|---|---|---|---|---|
| Mean | 615 | 690 | 163 | 5 | 60 | 310 |
| Median | 875 | 875 | 80 | 0 | 98 | 428 |
| Range | (0, 875) | (0, 875) | (2.5, 875) | (0, 30) | (0, 188) | (20, 875) |
| % Responder | 90% | 90% | 80% | 20% | 80% | 100% |

| ASC IgA | SE LPS | STm LPS | Vi | SE FliC | STm FliC | TT |
|---|---|---|---|---|---|---|
| Mean | 524 | 737 | 349 | 8 | 20 | 24 |
| Median | 491 | 875 | 229 | 4 | 13 | 16 |
| Range | (30, 875) | (60, 875) | (28, 875) | (0, 28) | (0, 60) | (0, 65) |
| % Responder | 100% | 100% | 100% | 30% | 60% | 60% |

Placebo, n=2

| ASC IgG | SE LPS | STm LPS | Vi | SE FliC | STm FliC | TT |
|---|---|---|---|---|---|---|
| Range | (0, 3) | (0, 0) | (0, 0) | (0, 0) | (0, 0) | (0, 0) |
| % Responder | 0 | 0 | 0 | 0 | 0 | 0 |

| ASC IgA | SE LPS | STm LPS | Vi | SE FliC | STm FliC | TT |
|---|---|---|---|---|---|---|
| Range | (0, 0) | (3, 5) | (0, 0) | (0, 0) | (0, 0) | (0, 0) |
| % Responder | 0 | 0 | 0 | 0 | 0 | 0 |

#### Table 5: Sorted Antibody Secreting Cells Responses, for Tissue Homing Potential

Sorted ASC responses were measured from Cohort B participants. Memory B cells (CD19+ CD27+) were sorted into 4 subpopulations: CD62L+ α4β7- (LN: Lymph Node homing), CD62L- α4β7- (unknown homing), CD62L+ α4β7+ (LN and gut mucosa homing), and CD62L- α4β7+ (gut mucosa homing) from recipients of 12.5μg of TSCV intramuscularly (n=10). Antigen-specific cells detected for each sorted subpopulation were converted to Spot Forming Cells (SFC)/million of purified CD19+CD27+ cells based on the number of sorted cells seeded in each well. Subjects with >500 SFC/million cells in at least 2 out of 4 sorted subsets for each antigen were included in the analysis.

| | | **62L-<br>A4β7-<br><br>(unknown homing)** | **62L+<br>A4β7-<br><br>(Lymph Node)** | **62L+<br>A4β7+<br>(LN AND Gut Mucosa)** | **62L-<br>A4β7+<br><br>(Gut Mucosa)** |
|---|---|---|---|---|---|
| **Vi IgG** | | | | | |
| | Mean | 2,156 | 6,723 | 746 | 851 |
| | Median | 1,135 | 2,727 | 833 | 0 |
| | Range | 0-8,250 | 58-32,432 | 0-1,429 | 0-5,641 |
| **Vi IgA** | | | | | |
| | Mean | 4,578 | 6,863 | 12,670 | 8,908 |
| | Median | 5,000 | 4,886 | 13,966 | 8,622 |
| | Range | 0-10,702 | 1,516-14,148 | 855-32,222 | 0-22,167 |
| **SE IgG** | | | | | |
| | Mean | 5,610 | 6,758 | 8,259 | 7,738 |
| | Median | 4,512 | 3,472 | 6,827 | 5,917 |
| | Range | 0-19,000 | 729-19,773 | 2,286-19,130 | 0-24,375 |
| **SE IgA** | | | | | |
| | Mean | 6,642 | 6,278 | 16,133 | 22,283 |
| | Median | 2,994 | 5,442 | 11,467 | 20,029 |
| | Range | 1,000-21,111 | 2,102-12,923 | 5,000-37,586 | 0-45,667 |
| **STm IgG** | | | | | |
| | Mean | 5,774 | 11,349 | 18,645 | 13,280 |
| | Median | 4,693 | 12,810 | 16,251 | 10,682 |
| | Range | 0-18,000 | 1,166-19,773 | 3,519-42,391 | 0-34,667 |
| **STm IgA** | | | | | |
| | Mean | 9,380 | 10,947 | 31,565 | 35,115 |
| | Median | 8,700 | 10,923 | 25,841 | 37,037 |
| | Range | 0-20,556 | 6,592-17,796 | 5,600-76,000 | 0-71,250 |

### Table 6. Memory B Cell Responses

Antigen-specific memory B ($B_M$) cell responses were measured and are reported by the % of specific $B_M$ per total B cells.

#### 6.25 µg TSCV

| $B_m$ IgG Day 1 | Vi | SE LPS | STm LPS | SE FliC | STm FliC | TT | SCh LPS |
|---|---|---|---|---|---|---|---|
| N | 8 | 8 | 8 | 7 | 7 | 7 | 7 |
| Mean | 0 | 0.0343 | 0.009 | 0.013 | 0.013 | 0.160 | 0 |
| Median | 0 | 0 | 0 | 0 | 0 | 0.110 | 0 |
| Range | 0, 0 | 0, 0.190 | 0, 0.07 | 0, 0.09 | 0, 0.9 | 0, 0.43 | 0, 0 |
| % Responder | 0% | 12.5% | 0% | 0% | 0% | 71.4% | 0% |

| $B_m$ IgA Day 1 | Vi | SE LPS | STm LPS | SE FliC | STm FliC | TT | SCh LPS |
|---|---|---|---|---|---|---|---|
| N | 8 | 8 | 8 | 7 | 7 | 7 | 7 |
| Mean | 0 | 0.106 | 0.009 | 0.071 | 0.131 | 0.047 | 0 |
| Median | 0 | 0 | 0 | 0 | 0 | 0 | 0 |
| Range | 0, 0 | 0, 0.57 | 0, 0.07 | 0, 0.50 | 0, 0.50 | 0, 0.33 | 0, 0 |
| % Responder | 0% | 25% | 0% | 14.3% | 28.6% | 14.3% | 0% |

| $B_m$ IgG Day 29 | Vi | SE LPS | STm LPS | SE FliC | STm FliC | TT | SCh LPS |
|---|---|---|---|---|---|---|---|
| N | 8 | 8 | 8 | 7 | 7 | 7 | 6 |
| Mean | 0.370 | 0.128 | 0.126 | 0.120 | 0.130 | 1.10 | 0.055 |
| Median | 0.285 | 0.085 | 0.085 | 0.070 | 0.130 | 0.89 | 0 |
| Range | 0, 1.08 | 0, 0.34 | 0, 0.33 | 0, 0.37 | 0, 0.32 | 0.12, 2.47 | 0, 0.295 |
| % Responder | 50% | 37.5% | 62.5% | 14.3% | 57.1% | 85.7% | 16.7% |

| $B_m$ IgA Day 29 | Vi | SE LPS | STm LPS | SE FliC | STm FliC | TT | SCh LPS |
|---|---|---|---|---|---|---|---|
| N | 8 | 8 | 8 | 7 | 7 | 7 | 5 |
| Mean | 1.63 | 0.648 | 0.126 | 7 | 0.123 | 0 | 0.042 |
| Median | 0.63 | 0.44 | 0.085 | 7 | 0 | 0 | 0 |
| Range | 0, 7.15 | 0, 1.81 | 0, 0.33 | 0, 0 | 0, 0.51 | 0, 0 | 0, 0.21 |
| % Responder | 75% | 75% | 37.5% | 0% | 14.3% | 0% | 20% |

| $B_m$ IgG Day 57 | Vi | SE LPS | STm LPS | SE FliC | STm FliC | TT | SCh LPS |
|---|---|---|---|---|---|---|---|
| N | 7 | 8 | 7 | 7 | 7 | 7 | 6 |
| Mean | 0.210 | 0.0338 | 0.093 | 0.029 | 0.086 | 0.81 | 0 |
| Median | 0.230 | 0 | 0.07 | 0 | 0 | 0.81 | 0 |
| Range | 0, 0.55 | 0, 0.17 | 0, 0.28 | 0, 0.20 | 0, 0.32 | 0.22, 1.39 | 0, 0 |
| % Responder | 57% | 12.5% | 42.9% | 28.6% | 28.6% | 87.5% | 0% |

| $B_m$ IgA Day 57 | Vi | SE LPS | STm LPS | SE FliC | STm FliC | TT | SCh LPS |
|---|---|---|---|---|---|---|---|
| N | 7 | 7 | 7 | 7 | 7 | 7 | 6 |
| Mean | 0.71 | 0.447 | 0.093 | 0 | 0 | 0 | 0 |
| Median | 0.41 | 0.56 | 0.07 | 0 | 0 | 0 | 0 |
| Range | 0, 2.06 | 0, 0.92 | 0, 0.28 | 0, 0 | 0, 0 | 0, 0 | 0, 0 |
| % Responder | 71.4% | 57.1% | 28.6% | 0% | 0% | 0% | 0% |

| $B_m$ IgG D510 | Vi | SE LPS | STm LPS | SE FliC | STm FliC | TT | SCh LPS |
|---|---|---|---|---|---|---|---|
| N | 7 | 7 | 7 | 6 | 6 | 6 | 5 |
| Mean | 0 | 0.027 | 0 | 0.055 | 0.020 | 0.242 | 0 |
| Median | 0 | 0 | 0 | 0 | 0 | 0.20 | 0 |
| Range | 0, 0 | 0, 0.19 | 0, 0 | 0, 0.33 | 0, 0.12 | 0.12, 0.53 | 0, 0 |
| % Responder | 0% | 0% | 0% | 16.7% | 16.7% | 50% | 0% |

| $B_m$ IgA D510 | Vi | SE LPS | STm LPS | SE FliC | STm FliC | TT | SCh LPS |
|---|---|---|---|---|---|---|---|
| N | 7 | 6 | 7 | 6 | 6 | 7 | 5 |
| Mean | 0.104 | 0.120 | 0 | 0 | 0 | 0 | 0 |
| Median | 0 | 0.085 | 0 | 0 | 0 | 0 | 0 |
| Range | 0, 0.40 | 0, 0.37 | 0, 0 | 0, 0 | 0, 0 | 0, 0 | 0, 0 |
| % Responder | 28.6% | 33.3% | 0% | 0% | 0% | 0% | 0% |

#### 12.5 µg TSCV

| $B_m$ IgG Day 1 | Vi | SE LPS | STm LPS | SE FliC | STm FliC | TT | SCh LPS |
|---|---|---|---|---|---|---|---|
| N | 10 | 10 | 10 | 10 | 10 | 10 | 10 |
| Mean | 0 | 0 | 0 | 0.02 | 0.012 | 0.143 | 0 |
| Median | 0 | 0 | 0 | 0 | 0 | 0.105 | 0 |
| Range | 0, 0 | 0, 0 | 0, 0 | 0, 0.16 | 0, 0.12 | 0, 0.4 | 0, 0 |
| % Responder | 0% | 0% | 0% | 10% | 10% | 50% | 0% |

| $B_m$ IgA Day 1 | Vi | SE LPS | STm LPS | SE FliC | STm FliC | TT | SCh LPS |
|---|---|---|---|---|---|---|---|
| N | 10 | 10 | 10 | 9 | 10 | 10 | 10 |
| Mean | 0.14 | 0.087 | 0.100 | 0 | 0 | 0.019 | 0 |
| Median | 0 | 0 | 0 | 0 | 0 | 0 | 0 |
| Range | 0, 0.69 | 0, 0.57 | 0, 0.56 | 0, 0 | 0, 0 | 0, 0.19 | 0, 0 |
| % Responder | 40% | 20% | 20% | 0% | 0% | 10% | 0% |

| $B_m$ IgG Day 29 | Vi | SE LPS | STm LPS | SE FliC | STm FliC | TT | SCh LPS |
|---|---|---|---|---|---|---|---|
| N | 8 | 10 | 8 | 10 | 9 | 8 | 10 |
| Mean | 0.19 | 0.059 | 0.10 | 0.009 | 0.078 | 0.55 | 0 |
| Median | 0.145 | 0 | 0 | 0 | 0 | 0.3 | 0 |
| Range | 0, 0.5 | 0, 0.38 | 0, 0.76 | 0, 0.09 | 0.49 | 0.07, 1.22 | 0, 0 |
| % Responder | 62.5% | 30% | 12.5% | 0% | 22.2% | 87.5% | 0% |

| $B_m$ IgA Day 29 | Vi | SE LPS | STm LPS | SE FliC | STm FliC | TT | SCh LPS |
|---|---|---|---|---|---|---|---|
| N | 8 | 10 | 8 | 10 | 9 | 7 | 10 |
| Mean | 1.135 | 0.344 | 0.703 | 0.097 | 0.064 | 0 | 0 |
| Median | 0.74 | 0.21 | 0.67 | 0 | 0 | 0 | 0 |
| Range | 0, 3.38 | 0, 107 | 0.16, 1.38 | 0, 0.74 | 0, 0.42 | 0, 0 | 0, 0 |
| % Responder | 75% | 50% | 100% | 20% | 22.2% | 0% | 0% |

| $B_m$ IgG Day 57 | Vi | SE LPS | STm LPS | SE FliC | STm FliC | TT | SCh LPS |
|---|---|---|---|---|---|---|---|
| N | ns | ns | ns | ns | ns | ns | ns |
| Mean | | | | | | | |
| Median | | | | | | | |
| Range | | | | | | | |
| % Responder | | | | | | | |

| $B_m$ IgA Day 57 | Vi | SE LPS | STm LPS | SE FliC | STm FliC | TT | SCh LPS |
|---|---|---|---|---|---|---|---|
| N | ns | ns | ns | ns | ns | ns | ns |
| Mean | | | | | | | |
| Median | | | | | | | |
| Range | | | | | | | |
| % Responder | | | | | | | |

| $B_m$ IgG D450 | Vi | SE LPS | STm LPS | SE FliC | STm FliC | TT | SCh LPS |
|---|---|---|---|---|---|---|---|
| N | 9 | 10 | 9 | 10 | 9 | 9 | 9 |
| Mean | 0.101 | 0 | 0 | 0.006 | 0.019 | 0.17 | 0 |
| Median | 0 | 0 | 0 | 0 | 0 | 0 | 0 |

| $B_m$ IgA D450 | Vi | SE LPS | STm LPS | SE FliC | STm FliC | TT | SCh LPS |
|---|---|---|---|---|---|---|---|
| N | 9 | 9 | 9 | 10 | 9 | 9 | 9 |
| Mean | 0.568 | 0.066 | 0.67 | 0.036 | 0 | 0 | 0 |
| Median | 0.35 | 0 | 0 | 0 | 0 | 0 | 0 |

| | Vi | SE LPS | STm LPS | SE FliC | STm FliC | TT | SCh LPS |
|---|---|---|---|---|---|---|---|
| Range | 0,1.88 | 0, 0.36 | 0, 0 | 0, 0.06 | 0, 0.17 | 0, 0.73 | 0, 0 |
| % Responder | 22.2% | 0% | 0% | 0% | 11.1% | 44.4% | 0% |

| | Vi | SE LPS | STm LPS | SE FliC | STm FliC | TT | SCh LPS |
|---|---|---|---|---|---|---|---|
| Range | 0, 1.88 | 0, 0.36 | 0, 0.6 | 0, 0.36 | 0, 0 | 0, 0 | 0, 0 |
| % Responder | 55.6% | 11.1% | 11.1% | 10% | 0% | 0% | 0% |

#### Placebo

| $B_m$ IgG Day 1 | Vi | SE LPS | STm LPS | SE FliC | STm FliC | TT | SCh LPS |
|---|---|---|---|---|---|---|---|
| N | 4 | 4 | 4 | 4 | 4 | 4 | 4 |
| Mean | 0 | 0 | 0 | 0.033 | 0.018 | 0.103 | 0 |
| Median | 0 | 0 | 0 | 0 | 0 | 0.055 | 0 |
| Range | 0, 0 | 0, 0 | 0, 0 | 0, 0.13 | 0, 0.07 | 0, 0.30 | 0, 0 |
| % Responder | 0% | 0% | 0% | 25% | 0% | 50% | 0% |

| $B_m$ IgA Day 1 | Vi | SE LPS | STm LPS | SE FliC | STm FliC | TT | SCh LPS |
|---|---|---|---|---|---|---|---|
| N | 4 | 4 | 4 | 4 | 4 | 4 | 4 |
| Mean | 0 | 0 | 0 | 0 | 0.018 | 0 | 0 |
| Median | 0 | 0 | 0 | 0 | 0 | 0 | 0 |
| Range | 0, 0 | 0, 0 | 0, 0 | 0, 0 | 0, 0.07 | 0, 0 | 0, 0 |
| % Responder | 0% | 0% | 0% | 0% | 0% | 0% | 0% |

| $B_m$ IgG Day 29 | Vi | SE LPS | STm LPS | SE FliC | STm FliC | TT | SCh LPS |
|---|---|---|---|---|---|---|---|
| N | 4 | 4 | 4 | 4 | 4 | 4 | 3 |
| Mean | 0 | 0 | 0.013 | 0 | 0.025 | 0.135 | 0.002 |
| Median | 0 | 0 | 0 | 0 | 0 | 0.125 | 0 |
| Range | 0, 0 | 0, 0 | 0, 0.05 | 0, 0 | 0, 0.1 | 0, 0.29 | 0, 0.004 |
| % Responder | 0% | 0% | 0% | 0% | 25% | 25% | 0% |

| $B_m$ IgA Day 29 | Vi | SE LPS | STm LPS | SE FliC | STm FliC | TT | SCh LPS |
|---|---|---|---|---|---|---|---|
| N | 4 | 4 | 4 | 4 | 4 | 4 | 4 |
| Mean | 0 | 0 | 0.073 | 0 | 0.025 | 0 | 0 |
| Median | 0 | 0 | 0 | 0 | 0 | 0 | 0 |
| Range | 0, 0 | 0, 0 | 0, 0.29 | 0, 0 | 0, 0.1 | 0, 0 | 0, 0 |
| % Responder | 0% | 0% | 25% | 0% | 0% | 0% | 0% |

| $B_m$ IgG Day 57 | Vi | SE LPS | STm LPS | SE FliC | STm FliC | TT | SCh LPS |
|---|---|---|---|---|---|---|---|
| N | 2 | ns | ns | 2 | 2 | 2 | 2 |
| Mean | 0 | | | 0 | 0.02 | 0.205 | 0 |
| Median | 0 | | | 0 | 0.02 | 0.205 | 0 |
| Range | 0, 0 | | | 0, 0 | 0, 0.04 | 0.2, 0.21 | 0, 0 |
| % Responder | 0% | | | 0% | 0% | 50% | 0% |

| $B_m$ IgA Day 57 | Vi | SE LPS | STm LPS | SE FliC | STm FliC | TT | SCh LPS |
|---|---|---|---|---|---|---|---|
| N | 2 | ns | ns | ns | ns | ns | 2 |
| Mean | 0.125 | | | | | | 0 |
| Median | 0.125 | | | | | | 0 |
| Range | 0, 0.25 | | | | | | 0, 0 |
| % Responder | 50% | | | | | | 0% |

| $B_m$ IgG D450/510 | Vi | SE LPS | STm LPS | SE FliC | STm FliC | TT | SCh LPS |
|---|---|---|---|---|---|---|---|
| N | 2 | 2 | 2 | 2 | 2 | 2 | 2 |
| Mean | 0 | 0 | 0.04 | 0 | 0 | 0.09 | 0 |
| Median | 0 | 0 | 0.04 | 0 | 0 | 0.09 | 0 |
| Range | 0, 0 | 0, 0 | 0, 0.08 | 0, 0 | 0, 0 | 0, 0.18 | 0, 0 |
| % Responder | 0% | 0% | 0% | 0% | 0% | 50% | 0% |

| $B_m$ IgA D450/510 | Vi | SE LPS | STm LPS | SE FliC | STm FliC | TT | SCh LPS |
|---|---|---|---|---|---|---|---|
| N | 2 | 2 | 2 | 4 | 4 | 4 | 2 |
| Mean | 0 | 0 | 0 | 0 | 0.05 | 0 | 0 |
| Median | 0 | 0 | 0 | 0 | 0 | 0 | 0 |
| Range | 0, 0 | 0, 0 | 0, 0 | 0, 0 | 0, 0.2 | 0, 0 | 0, 0 |
| % Responder | 0% | 0% | 0% | 0% | 25% | 0% | 0% |

ns, no samples

<u>Antigens</u>:
Vi = *S*. Typhi Vi Polysaccharide
SE LPS = *S*. Enteritidis Lipopolysaccharide
STm LPS = *S*. Typhimurium Lipopolysaccharide
SE FliC = *S*. Enteritidis FliC flagellin subunit
STm FliC = *S*. Typhimurium FliC flagellin subunit
TT = tetanus toxoid (positive control)
SCh LPS = *S*. Choleraesuis Lipopolysaccharide (negative control)

### Figure 7. Gating Protocol of Sorted B cell subpopulations

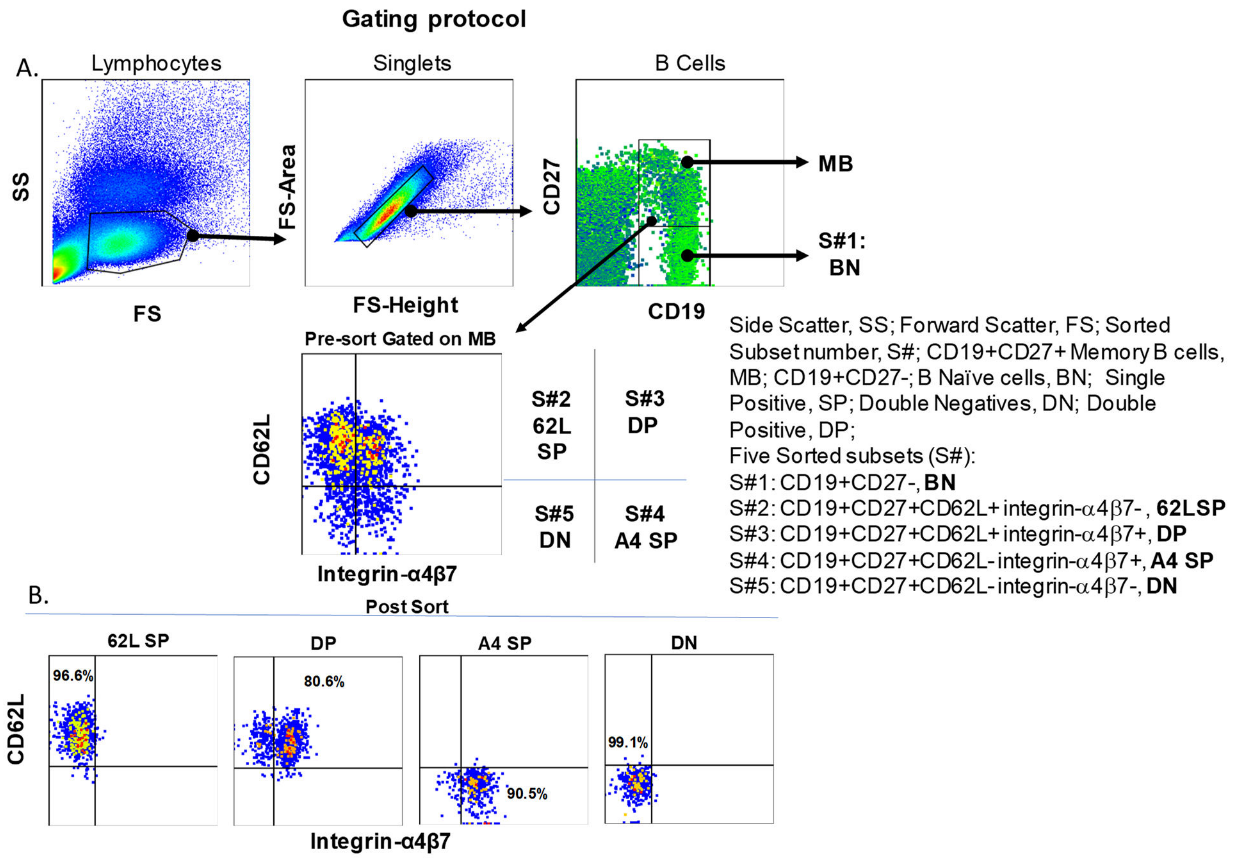

**Representative gating protocol for B cell sorting.** Freshly isolated PBMC purified from subjects 7-days after vaccination with TSCV (Day 8) were stained with monoclonal antibodies against CD19, CD27, CD62L, and integrin α4β7 markers and the stained cells were simultaneously sorted into five different B cell subsets, using a MoFlow Astrios cell sorter (Beckman-Coulter) as described in the methods section. Sequential gating protocols are shown in **panel A**. An aliquot of B Memory (BM) cell sorted populations was analyzed post-sorting to assess purity of the subsets **(panel B).**

#### Figure 8. Percentage of antigen-specific BM responses

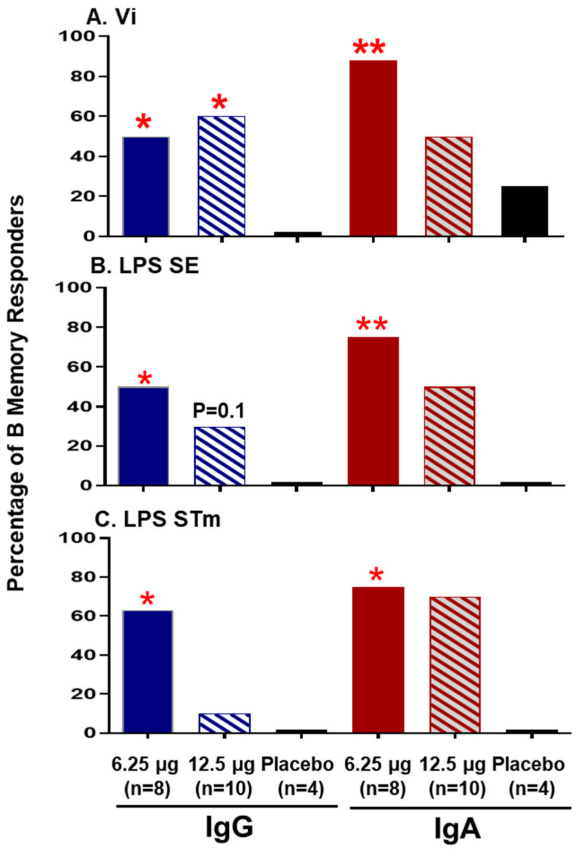

**Percentages of responders showing increased antigen-specific B memory responses to polysaccharide antigens in TSCV following vaccination.** Shown are the percentages of responders for the induction of IgG and IgA BM responses specific for Vi from *S*. Typhi (**panel A**), LPS from *S*. Enteritidis - LPS SE (panel B) and LPS from *S*. Typhimurium - STm (**panel C**), following immunization with 6.25 µg or 12.5 µg TSCV or Placebo. A cutoff value for antigen specific responses was calculated as the Mean+3 SE of the corresponding antigen specific IgG and IgA BM responses in pre-vaccination (D1) samples from all participants (n=22). Volunteers showing post-vaccination increases above the corresponding antigen specific pre-vaccination cut-off values at any of the post-vaccination time points (Day 29, Day 57 or days 450/510) were considered responders. P values were determined by comparing vaccinated volunteers with the corresponding antigens in the placebo group (n=4) using Chi square tests. *p<0.05, ** p<0.01.
